# Odour cueing during sleep augments trauma-focused psychotherapy

**DOI:** 10.64898/2026.09.14.26362979

**Authors:** Anja Schaich, Sabine Groch, Mojgan Ehsanifard, Jovana Lehmann-Grube, Clara Sayk, Hong-Viet V Ngo-Dehning, Mathias Kammerer, Eva Fassbinder, Judith Amores, Frieder Paulus, Sören Krach, Jan-Philipp Klein, Ines Wilhelm

## Abstract

Trauma-focused psychotherapies reduce posttraumatic stress disorder (PTSD) symptoms by updating maladaptive traumatic memories into more adaptive representations. However, many patients do not achieve clinically meaningful improvement, highlighting the need for augmentation strategies. Targeted memory reactivation (TMR), in which memory cues are re-presented during sleep to strengthen memory consolidation, is a promising approach. We combined a mechanistic study in healthy participants with a randomized clinical trial to test whether odour cueing during sleep augments trauma-focused psychotherapy. In healthy participants, re-exposure to the therapy-associated odour during sleep increased slow-wave activity and slow oscillations, while greater increases in slow oscillation–spindle coupling were associated with reduced emotional arousal during recall of aversive autobiographical memories. In patients with PTSD, repeated odour cueing during sleep throughout 12 weeks of trauma-focused treatment improved clinician-rated and self-reported PTSD symptoms, particularly re-experiencing and negative alterations in cognition and mood, and accelerated treatment response and remission. These findings provide converging mechanistic and clinical evidence that TMR can enhance trauma-focused psychotherapy, potentially through sleep-dependent memory consolidation.

## Introduction

Post-traumatic stress disorder (PTSD) is characterized by maladaptive processing of traumatic memories, resulting in intrusive re-experiencing, avoidance, hyperarousal, and substantial functional impairment^1^. In addition to severe psychological distress, PTSD is associated with increased rates of comorbid mental and physical disorders, impaired psychosocial functioning, and reduced quality of life^2–5^. The disorder also imposes considerable societal costs through increased healthcare utilization and productivity losses^6^. Trauma-focused psychotherapies aim to alleviate PTSD symptoms by promoting adaptive reprocessing of traumatic memories and thereby transforming maladaptive memory representations into more adaptive representations^7^. Although these treatments are effective for many patients, approximately 40% do not achieve a clinically meaningful response and around 21% discontinue treatment prematurely^8,9^. This highlights the need for augmentation strategies that strengthen therapy-related learning beyond the therapy session and thereby improve treatment outcomes.

Sleep provides a mechanistically plausible target to support therapeutic change^10–12^. Recent findings indicate that sleep facilitates both the consolidation of newly acquired information and the reconsolidation of previously stored memories following updating^13–15^. In particular, slow oscillations, sleep spindles, and their precise temporal coupling have been shown to support the stabilization and long-term integration of memories^16,17^. Building on the idea that spontaneous memory reactivation during sleep is a key mechanism underlying sleep-dependent memory consolidation, targeted memory reactivation (TMR) was developed to externally enhance these endogenous reactivation processes. In TMR, sensory cues such as sounds or odours are paired with recently encoded experiences and subsequently re-presented during sleep to promote memory reactivation and thereby strengthen consolidation and subsequent retrieval^18–21^.

Although TMR reliably enhances memory performance in laboratory settings, its translation into clinical interventions remains limited. Initial studies combining TMR with brief exposure-based interventions for specific phobia and social anxiety disorder failed to demonstrate additional clinical benefits beyond those achieved by the intervention alone^22,23^. Evidence for trauma-related disorders is even more limited. In healthy participants, repeated nights of TMR following an imagery rescripting intervention, in which aversive autobiographical memories are modified into more adaptive representations through mental imagery, further reduced vividness, distress, and emotional arousal associated with the updated memory^24^. Likewise, a recent study in patients with PTSD reported that TMR following a single session of Eye Movement Desensitization and Reprocessing therapy enhanced slow oscillatory and spindle activity and showed that these changes were associated with symptom improvement^25^. However, the absence of an effect on overall PTSD symptom severity limits conclusions regarding the clinical effectiveness of TMR. Thus, it remains unclear whether sleep-based memory reactivation can meaningfully enhance the outcomes of established trauma-focused psychotherapies and which neurophysiological mechanisms may underlie such effects.

Here, we tested whether TMR using odour cueing can augment trauma-focused interventions that aim to update aversive autobiographical memories. To address both mechanistic and clinical questions, we conducted two complementary studies. In the mechanistic study, we examined the behavioural and neural effects of odour cueing during sleep following a single imagery-based memory updating intervention in healthy participants. Neural mechanisms were assessed using high-density electroencephalography (EEG) in a within-subject design. In the clinical study, we tested the effect of repeated odour cueing during a full course of trauma-focused psychotherapy in a double-blind randomized controlled trial of patients with PTSD. In both studies, aversive memories were updated in the presence of an odour cue that was subsequently re-presented during sleep to induce memory reactivation, whereas control participants were exposed to a different odour.

We show that nocturnal memory reactivation using odour cues enhances sleep oscillatory activity implicated in memory consolidation, particularly slow-wave activity and slow oscillation–spindle coupling. Furthermore, greater increases in slow oscillation–spindle coupling predicted larger reductions in emotional arousal of updated memories. Furthermore, repeated odour cueing during sleep improves clinically relevant outcomes of trauma-focused psychotherapy in PTSD by accelerating treatment response and remission and by reducing core PTSD symptoms. Together, these findings provide mechanistic and clinical evidence that sleep-dependent memory reactivation using odour cueing can strengthen the consolidation of adaptive memory representations formed during trauma-focused psychotherapy.

## Results

### Study 1: Mechanistic evidence for sleep-related reactivation of updated autobiographical memories

Twenty-eight healthy university students (24 women; mean age 23.36 ± 3.29 years) participated in Study 1. Participants provided two negative and two neutral autobiographical memories, which were transformed into individualized audio scripts for script-driven imagery, a procedure commonly used to assess emotional responses to autobiographical memories^26^. In a within-subject design, one negative and one neutral memory were assigned to each of two experimental conditions (Fig. 1a). Script-driven imagery assessments were conducted before the intervention (T0), the morning after sleep (T1), and again seven days later (T2; Fig. 1a,b). Following baseline script-driven imagery (T0), participants received one session of imagery rescripting and reprocessing therapy (IRRT), a trauma-focused intervention that updates aversive autobiographical memories through mental rescripting^27^. During IRRT, an odour cue was presented to become associated with the updated autobiographical memory. During subsequent sleep, high-density EEG was recorded while participants were re-exposed either to the odour previously presented during IRRT (congruent condition) or to a novel odour (incongruent condition; Fig. 1c).

**Figure 1.**
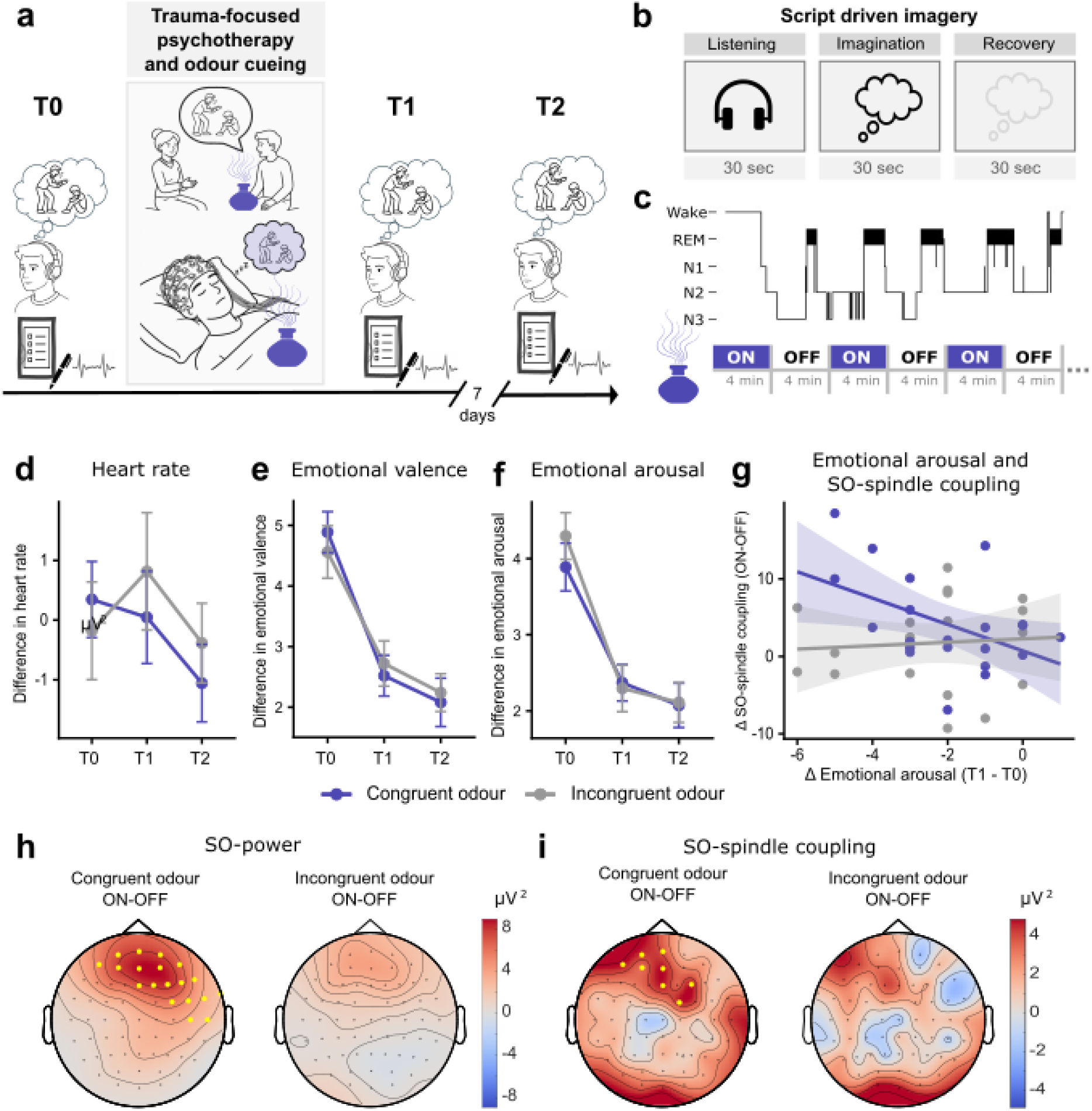
Study design and behavioural, physiological, and sleep EEG outcomes of Study 1. **(a)** Schematic overview of the study design. In the evening, participants first completed script-driven imagery (SDI; T0), during which emotional responses to individualized negative and neutral autobiographical memories were assessed using ratings of emotional arousal, valence, and vividness, as well as continuous heart rate recordings. This was followed by one session of imagery rescripting and reprocessing therapy (IRRT) targeting the negative autobiographical memory in the presence of an odour cue. During subsequent sleep, participants were re-exposed either to the same odour presented during IRRT (congruent condition) or to a novel odour (incongruent condition). Only the congruent condition is illustrated. SDI was repeated the following morning (T1) and 7 days later (T2). **(b)** Procedure of script-driven imagery (SDI). Each SDI session consisted of 30-s blocks of listening to an individualized audio script, imagining the autobiographical memory, and recovery. **(c)** Schematic illustration of nocturnal odour cueing during sleep with alternating 4-min odour ON and OFF periods. **(d)** Heart rate difference scores (negative − neutral memory) during SDI across assessment time points (T0, T1, T2). Heart rate responses did not change across assessments. **(e,f)** Emotional arousal and negative emotional valence (negative − neutral difference scores) during SDI across assessment time points. Emotional arousal and negative emotional valence decreased significantly following IRRT. **(g)** Association between SO–spindle coupling (averaged across electrodes within the significant cluster shown in h) and changes in emotional arousal (T1 − T0) in the congruent (blue) and incongruent (grey) conditions. In the congruent condition, greater SO–spindle coupling was associated with larger reductions in emotional arousal during negative SDI, whereas no significant association was observed in the incongruent condition. **(h)** Effects of nocturnal odour cueing on slow oscillations (SO) and **(i)** SO–spindle coupling. Topographic maps show odour ON versus OFF differences during congruent (left) and incongruent (right) cueing. Significant electrode clusters (*P* < 0.05) are highlighted in yellow.

#### Odour cueing does not further enhance behavioural responses following imagery rescripting and reprocessing therapy

To determine whether odour cueing enhances the effects of IRRT, we assessed subjective emotional responses and physiological reactivity during script-driven imagery of the neutral and negative autobiographical memory scripts. All outcome measures were analysed as negative–neutral difference scores. Emotional arousal and negative emotional valence decreased following IRRT (main effect of time: arousal, F(2,25) = 35.97, *P* < 0.001, η²ₚ = 0.74; negative emotional valence, F(2,23) = 32.97, *P* < 0.001, η²ₚ = 0.74; Fig. 1e,f), whereas neither vividness ratings (main effect of time: F(2,23) = 0.55, *P* = 0.583) nor heart rate responses (main effect of time: F(2,21) = 1.44, *P* = 0.259; Fig. 1d) changed across assessments. Importantly, odour cueing did not affect any subjective or physiological outcome measure, as indicated by the absence of significant condition effects or condition × time interactions (all *p* ≥ 0.149 for subjective ratings; all *p* ≥ 0.448 for heart rate; see Supplementary Tables S1, S2 for test statistics and descriptives). These findings indicate that IRRT reduced emotional responses to the autobiographical memory, whereas odour cueing did not provide additional behavioral benefits at the group level.

#### Odour cueing enhances sleep oscillatory signatures of memory consolidation

To identify neural correlates of odour cueing during sleep, we compared odour ON and OFF periods for oscillatory measures which were previously implicated in sleep-dependent memory consolidation, including slow oscillations (SO; 0.5–1.25 Hz), slow-wave activity (SWA; 0.5–4 Hz), fast spindle activity (12–16 Hz), SO–spindle coupling during non-rapid eye movement (NREM) sleep, and theta activity (4.25–8 Hz) during rapid eye movement (REM) sleep. Cluster-based permutation analyses of odour ON versus OFF periods revealed significantly increased frontal SO and SWA during congruent cueing, whereas no significant clusters emerged during incongruent cueing (Fig. 1h for SO, Supplementary Fig. S1 for SWA). Similarly, the absolute number of SO–spindle coupling events was significantly higher during odour ON than OFF periods in the congruent condition within a frontal electrode cluster, whereas no significant clusters were observed in the incongruent condition (Fig. 1i). No significant cueing effects were detected for fast spindle activity, REM theta activity, or overall sleep architecture (Supplementary Table S3). Together, these findings indicate that odour cueing selectively enhanced sleep oscillatory signatures implicated in memory consolidation.

#### Slow oscillation–spindle coupling is associated with behavioural responses following imagery rescripting and reprocessing therapy

Next, we examined whether the cue-related oscillatory changes identified above were associated with behavioural outcomes. Mean values from the significant SO, SWA and SO– spindle coupling clusters were correlated with behavioural measures of intervention success. Greater increases in SO–spindle coupling during congruent odour presentation were associated with larger reductions in emotional arousal during script-driven imagery (r(21) = −0.47, *p* = 0.031; Fig. 1g). No corresponding association was observed in the incongruent condition, and no significant correlations emerged for emotional valence, vividness, heart rate responses, SWA or SO (all *p* > 0.06; Supplementary Table S4).

### Study 2: Clinical effect of odour cueing in trauma-focussed therapy of post-traumatic stress disorder

Thirty-eight patients with PTSD (35 women; mean age, 38.0 ± 10.96 years) completed Study 2 (see Supplementary Fig. S2 for the CONSORT flow diagram). Patients were severely affected and had experienced at least one traumatic event before the age of 16 years, with many reporting multiple traumatic experiences. Sample characteristics and treatment information are summarized in Supplementary Table S5. Patients received 12 weekly sessions of imagery rescripting (ImRs), an imagery-based intervention that aims to transform maladaptive trauma-memory representations into more adaptive representations28. From the second session onwards, an odour cue was presented during therapy sessions and subsequently re-presented during sleep using a home diffuser (Fig. 2a). Patients were randomized to receive either the same odour previously presented during ImRs (congruent condition) or a different odour (incongruent condition). Clinician-rated and self-reported PTSD symptoms were assessed at baseline (T0), 1-month follow-up (T1), and 6-month follow-up (T2), with additional weekly assessments of self-reported PTSD symptoms throughout treatment. A subset of patients additionally underwent overnight sleep laboratory assessments at baseline and 1-month follow-up (N = 22).

**Figure 2.**
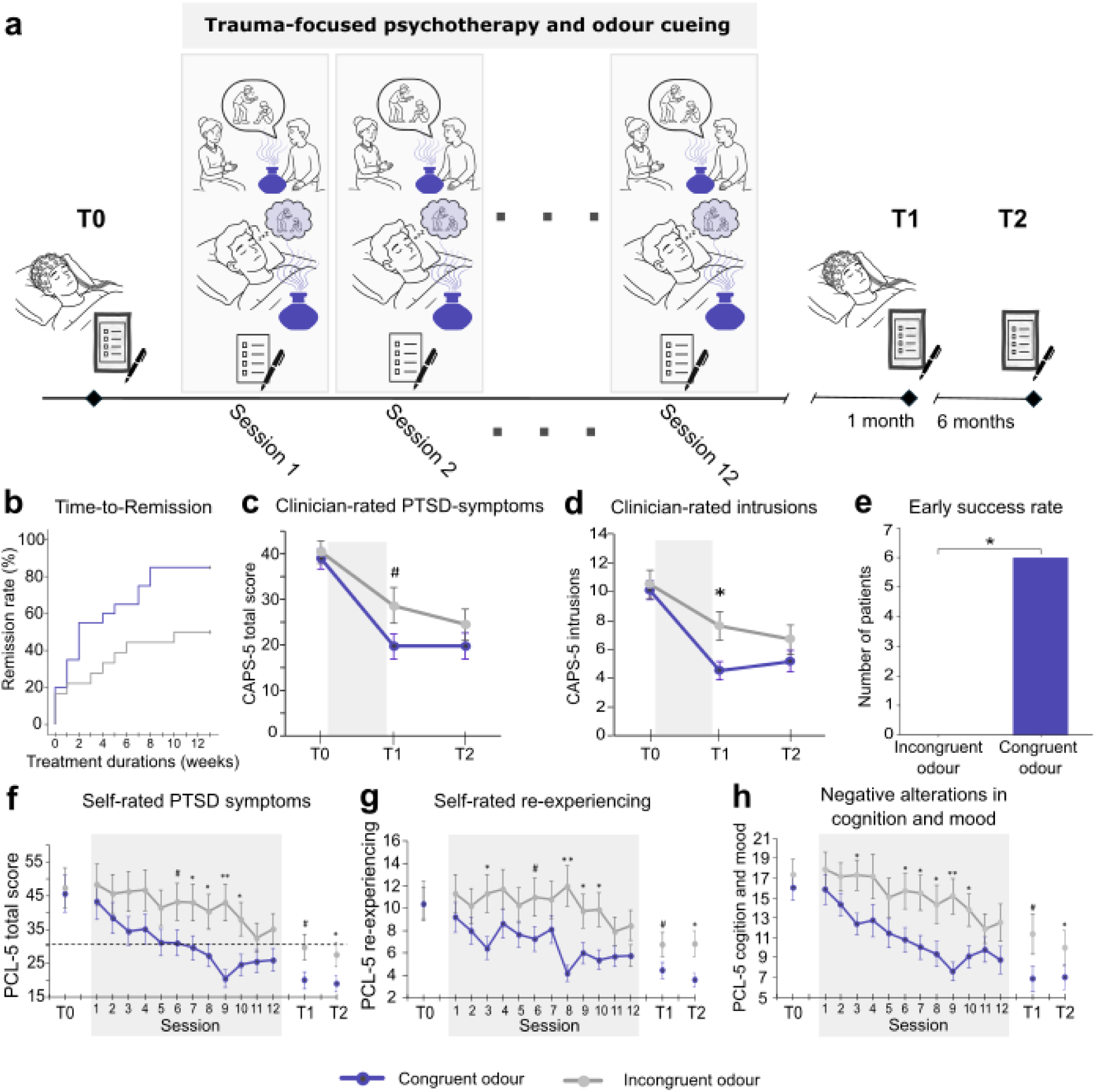
Study design and clinical outcomes of Study 2. **(a)** Schematic overview of the study design. Patients with PTSD were randomized to receive either the therapy-associated odour (congruent condition) or a different odour (incongruent condition) during sleep following 12 weekly sessions of imagery rescripting (ImRs). Only the congruent condition is illustrated. Clinical assessments were conducted at baseline (T0), 1-month follow-up (T1), and 6-month follow-up (T2). Overnight sleep laboratory assessments were additionally conducted at T0 and T1. **(b)** Kaplan–Meier estimates of time to remission derived from weekly assessments of self-reported PTSD symptoms assessed with PTSD Checklist for DSM-5 (PCL-5). Patients in the congruent condition (blue line) reached remission significantly earlier than patients in the incongruent condition (grey line). **(c,d)** Clinician-rated PTSD symptoms assessed with the Clinician-Administered PTSD Scale for DSM-5 (CAPS-5). Overall clinician-rated PTSD symptom severity showed a trend towards greater reductions in the congruent than in the incongruent condition. Clinician-rated intrusion symptoms decreased significantly more in the congruent than in the incongruent condition. **(e)** Early treatment success. Six patients (30%) in the congruent condition (blue bar) completed treatment before session 12 because remission criteria had been met, whereas no patients in the incongruent condition achieved early treatment success. **(f–h)** Greater reductions in self-reported PTSD symptoms were observed in the congruent (blue line) than in the incongruent condition (grey line) for the total score, the re-experiencing symptom cluster, and the negative alterations in cognition and mood symptom cluster. Data in panels f–h are shown as estimated marginal means ± s.e.m.

#### Odour cueing improves clinician-rated intrusion symptoms

Clinician-rated PTSD symptoms as assessed with the Clinician-Administered PTSD Scale for DSM-5 (CAPS-5^29^) decreased substantially over the course of treatment irrespective of cueing condition (main effect of time for CAPS-5 total score: *F*(2,32) = 26.06, *p* < 0.001; Supplementary Table S6). Although overall clinician-rated PTSD severity tended to decrease more strongly in the congruent than in the incongruent condition, this effect did not reach statistical significance (condition × time interaction: *F*(2,32) = 2.77, *p* = 0.077).

Analysis of CAPS-5 symptom clusters revealed that intrusion symptoms decreased more strongly in the congruent than in the incongruent odour-cueing condition (condition × time interaction: *F*(2,34) = 3.40, *p* = 0.045). Similar trends were observed for negative alterations in cognition and mood (*F*(2,41) = 2.52, *p* = 0.093) and hyperarousal symptoms (*F*(2,33) = 2.84, *p* = 0.073). Across both conditions, intrusion, avoidance, and negative alterations in cognition and mood symptoms decreased significantly over time (all *p* < 0.001), whereas dissociative symptoms showed a trend-level reduction (*p* = 0.060). Mean symptom trajectories are shown in Fig. 2 c,d; descriptive statistics, effect sizes, and full model results are provided in Supplementary Tables S6 and S7.

#### Odour cueing accelerates remission and treatment response

Patients that received congruent odour cueing reached remission faster than patients that received incongruent odour cueing (log-rank *χ²*(1) = 4.77, *p* = 0.029; Fig. 2b). Remission rates were 85% in the congruent condition and 50% in the incongruent condition, with estimated times to remission of 4.45 weeks (95% CI, 2.50–6.40) and 8.11 weeks (95% CI, 5.62–10.60), respectively. Congruent odour cueing also increased the likelihood of early treatment success. Six patients (30%) in the congruent condition completed treatment early, whereas no patients in the incongruent condition met criteria for early success (*χ²*(1) = 6.41, exact *p* = 0.021; Fig. 2e). Similarly, clinically meaningful improvement according to the minimal clinically important difference criterion (MCID) occurred more frequently in the congruent condition at mid-treatment (*χ²* = 9.75, *p* = 0.002; Supplementary Table S8). Additional analyses of treatment response and recovery are reported in the Supplementary Information (Supplementary Fig. S3).

#### Odour cueing improves self-reported posttraumatic stress symptoms

Self-reported PTSD symptoms as assessed with the Posttraumatic Stress Disorder Checklist for DSM-5 (PCL-5^30^) decreased across treatment in both conditions (main effect of time for PCL-5 total score: *F*(15,47) = 13.08, *p* < 0.001). Importantly, symptom reduction was greater in the congruent than in the incongruent condition (condition × time interaction: *F*(15,47) = 1.88, *p* = 0.024; Fig. 2f). Analysis of PCL-5 symptom clusters showed stronger reductions in re-experiencing symptoms (condition × time interaction: *F*(15,32) = 2.43, *p* = 0.002; Fig. 2g) and negative alterations in cognition and mood (*F*(15,13) = 1.91, *p* = 0.028; Fig. 2h) in the congruent condition. Descriptive statistics, effect sizes, and full model results are reported in Supplementary Tables S6 and S7. Post-traumatic cognitions, depressive symptoms, and anxiety symptoms decreased over the course of treatment, whereas self-efficacy and general functioning improved (all main effects of time *p* < 0.041). However, these improvements did not differ between cueing conditions, as indicated by the absence of significant condition × time interactions (all *p* > 0.176). Full results are provided in Supplementary Tables S6 and S7.

#### Sleep measures are largely unaffected by TMR

Subjective sleep quality improved across treatment irrespective of cueing condition (main effect of time: F(2,69) = 12.79, *p* < 0.001), with no evidence for differential changes between groups (condition × time interaction: F(2,69) = 0.39, *p* = 0.678). In the polysomnography subsample (N = 22), objective sleep architecture remained largely unchanged (Supplementary Table S6, S7). The only significant effect was a greater reduction in slow-wave sleep in the incongruent condition (F(2,6919) = 4.73, *p* = 0.042). Likewise, analyses of sleep oscillatory activity revealed no significant effects of condition, time, or their interaction for slow-wave activity, slow oscillations, or fast spindle activity (all *p* > 0.120; Supplementary Table S9).

## Discussion

Across two complementary studies, we investigated whether targeted memory reactivation (TMR) using odour cueing during sleep can augment trauma-focused psychotherapy by strengthening sleep-dependent consolidation of updated aversive memories. In healthy participants, nocturnal cueing following imagery rescripting and reprocessing therapy (IRRT) enhanced sleep oscillatory activity implicated in memory consolidation, including slow-wave activity (SWA), slow oscillation (SO), and slow oscillation (SO)–spindle coupling. Although these neural effects were not accompanied by a robust overall behavioural advantage, increases in SO–spindle coupling predicted reductions in emotional arousal of the rescripted memory. Extending these mechanistic findings to a clinical context, repeated home-based cueing during sleep throughout a full course of imagery rescripting (ImRs) treatment improved clinician-rated and self-reported PTSD symptoms, accelerated remission, and increased early treatment response in patients with PTSD. Together, these findings provide converging mechanistic and clinical evidence that odour cueing can enhance trauma-focused interventions and support the notion that sleep-dependent memory processes contribute to the long-term consolidation of adaptive memory representations formed during psychotherapy^12,31^.

In Study 1, nocturnal memory cueing modulated sleep oscillatory activity following IRRT. Using high-density EEG and cluster-based permutation analyses, we observed significant increases in SWA, SO and SO–spindle coupling within frontal electrode clusters during presentation of the congruent odour, whereas no significant clusters emerged during incongruent odour presentation. Moreover, increases in SO–spindle coupling during congruent cueing predicted greater reductions in emotional arousal of the rescripted memory. Together, these findings provide mechanistic evidence that sleep-based cueing can influence neural processes implicated in the consolidation of updated autobiographical memories.

Our findings on SO–spindle coupling align with growing evidence that memory consolidation depends not only on the occurrence of slow oscillations and sleep spindles, but also on their precise temporal coordination^32^. SO–spindle coupling has been linked to successful memory consolidation, memory reactivation strength, and systems-level redistribution of newly encoded information^33–35^. More broadly, hierarchical nesting of sleep oscillations is thought to create temporal windows that facilitate hippocampo-cortical communication and cortical plasticity during memory consolidation. Extending this framework to psychotherapy, our findings suggest that sleep-dependent coordination of oscillatory events may contribute to the consolidation of adaptive memory representations formed during trauma-focused interventions.

Despite these oscillatory effects and their association with individual differences in emotional memory responses, congruent cueing did not produce a significant behavioural advantage at the group level in Study 1. Several factors may account for this discrepancy. Participants were healthy young adults who recalled only moderately aversive autobiographical memories, and IRRT itself produced substantial reductions in emotional arousal and negative valence. Under these conditions, a single night of odour cueing may have provided limited additional behavioural benefit despite measurable modulation of sleep-dependent consolidation processes. This interpretation is broadly consistent with previous studies combining TMR with exposure-based interventions, in which exposure therapy alone substantially reduced symptoms whereas additional sleep cueing did not further enhance clinical outcomes^22,23^. In contrast, Recher et al.^24^ reported reductions in arousal, vividness, and distress following TMR of rescripted autobiographical memories in healthy individuals, potentially because participants received repeated nights of cueing rather than a single night of reactivation. Together, these findings suggest that the behavioural expression of TMR effects depends on several boundary conditions, including the amount of cueing, the strength and salience of pre-sleep learning, and the emotional relevance of the targeted memory^36^. Consistent with this interpretation, behavioural benefits of TMR became apparent in Study 2, in which odour cueing was repeatedly applied during treatment of clinically significant traumatic memories in patients with PTSD.

In Study 2, we translated the mechanistic findings from Study 1 into a clinical setting and examined whether repeated odour cueing during sleep could enhance the effects of imagery rescripting in patients with PTSD. To our knowledge, this is the first trial to combine odour cueing with a full course of trauma-focused psychotherapy in a clinical PTSD sample. Re-exposure to therapy-associated odours during sleep accelerated remission, increased early treatment response, and improved self-reported PTSD symptoms. In addition, clinician-rated improvements were observed for intrusion symptoms, a symptom domain closely linked to maladaptive trauma-memory processing. Together, these findings suggest that repeated odour cueing during sleep can enhance clinically relevant aspects of trauma-focused treatment.

Interestingly, effects of odour cueing were more pronounced for self-reported symptoms specifically related to the treated index trauma than for global clinician-rated PTSD severity. One possible explanation is that the CAPS-5 captures symptom burden associated with multiple traumatic experiences over the preceding month, whereas the PCL-5 assessed symptoms related to the index trauma during the previous week. Therapists were instructed to target the index trauma until session five. However, patients in our study were severely affected and commonly reported multiple traumatic experiences, the enhancement of memory consolidation processes through odour cueing may therefore have been most detectable for the index trauma which was addressed in any case in the first half of the treatment.

Notably, the most clinically relevant effect of odour cueing may not be an increase in final treatment outcome per se, but rather an acceleration of therapeutic improvement. Patients in the congruent condition reached remission earlier and were more likely to achieve early treatment success. Faster therapeutic response is highly relevant for patients because it reduces suffering sooner and may lower the risk of treatment disengagement. From a health-care perspective, accelerating treatment response could also contribute to shorter treatment durations and reduced treatment costs.

Several aspects of the present work strengthen the interpretation and translational relevance of the findings. Most importantly, both studies employed an active control condition in which participants were exposed to an odour during sleep that had not been associated with the therapeutic intervention. Because odours themselves can influence sleep physiology, including processes implicated in memory consolidation, this design provided a particularly stringent test of memory-specific reactivation effects. In contrast to sham or no-stimulation control conditions frequently used in previous TMR studies^22,25^, both groups received equivalent nocturnal sensory stimulation, differing only in whether the cue had been associated with therapeutic memory updating. A second strength lies in the combination of a mechanistic laboratory study with a randomized controlled clinical trial. While Study 1 provided evidence that odour cueing updated aversive memories during sleep by modulating oscillatory processes that are implicated in memory consolidation, Study 2 demonstrated clinically meaningful benefits of repeated nocturnal odour cueing during a full course of trauma-focused psychotherapy. The convergence of mechanistic and clinical findings strengthens the interpretation that sleep-based memory reactivation contributed to the observed treatment effects. Finally, the clinical trial was conducted in a severely affected PTSD sample characterized by early, often multiple traumatic experiences and it evaluated odour cueing as an adjunct to an established trauma-focused psychotherapy delivered under routine clinical conditions. Together with blinded clinician ratings, blinded therapists’ repeated symptom assessments, and weekly monitoring of treatment trajectories, this design increases the ecological validity and potential clinical relevance of the findings.

Several limitations should be considered. First, the clinical sample was modest in size, and only a subset of patients participated in sleep laboratory assessments, limiting statistical power for electrophysiological analyses. Second, the mechanistic and clinical findings were obtained in separate studies. Although Study 1 demonstrated cue-related modulation of sleep oscillatory activity, electrophysiological correlates of memory reactivation has not been assessed in the clinical trial. Consequently, it remains unclear whether the clinical benefits observed in Study 2 were mediated by the same sleep-dependent mechanisms identified in Study 1. Future studies combining sleep recordings with odour cueing during trauma-focussed psychotherapy may help to clarify the mechanisms and optimize the effectiveness of sleep-based memory reactivation.

In conclusion, the present findings provide converging mechanistic and clinical evidence that odour cueing during sleep can augment trauma-focused psychotherapy. Across two complementary studies, odour cueing modulated sleep oscillatory activity implicated in memory consolidation and improved clinically relevant outcomes in PTSD. Together, these findings suggest that adaptive memory representations formed during psychotherapy remain amenable to sleep-dependent processing and can be selectively strengthened through memory cueing during sleep. More broadly, our results identify sleep-dependent memory consolidation as a promising target for enhancing psychological treatments and highlight the potential of low-cost and scalable sleep-based interventions to improve mental health outcomes.

## Methods

### STUDY 1

*Participants* were eligible for Study 1 if they were (1) between 18 and 30 years of age and (2) reported at least two personally distressing autobiographical memories that were at least 2 years old. Exclusion criteria were (1) traumatic memories requiring clinical treatment, (2) current treatment for a psychological disorder, (3) presence of a sleep disorder or any condition that could interfere with polysomnographic recordings, (4) travel across two or more time zones or engagement in shift work within the past 30 days, (5) use of sleep-altering medication or substances, including daily alcohol consumption exceeding 60 g in men and 40 g in women, and (6) allergies, including skin allergies, hay fever, or olfactory disorders. For information on randomization and masking as well as sample size calculation please refer to the Supplementary Methods.

#### Sleep macrostructure and sleep EEG oscillatory activity

Sleep was recorded using 64-channel EEG together with electrooculography (EOG) and electromyography (EMG) (Brain Products GmbH, Gilching, Germany). Data were preprocessed using BrainVision Analyzer (version 2.2.0.7383) and MATLAB R2021b (MathWorks, Natick, MA, USA). Preprocessing included visual inspection, identification of bad channels and artefacts, interpolation of noisy channels, re-referencing to the averaged mastoids, and band-pass filtering between 0.03 and 35 Hz. Sleep stages were scored independently by two experienced raters blinded to experimental condition according to American Academy of Sleep Medicine (AASM) criteria. Spectral and event-based analyses were performed using custom MATLAB scripts based on the FieldTrip toolbox. Analyses focused on artefact- and arousal-free NREM and REM sleep epochs. To assess cueing-related oscillatory activity, EEG data were segmented into alternating 4-min odour ON and 4-min odour OFF intervals. Based on previous TMR studies, the first minute of each odour ON interval and the final minute of the preceding odour OFF interval were selected for analysis. Spectral power was estimated using Fast Fourier Transformation and averaged within predefined frequency bands: slow-wave activity (SWA; 0.75–4 Hz), fast spindle activity (12– 16 Hz) during NREM sleep, and theta activity (4.25–8 Hz) during REM sleep. To quantify SO– spindle coupling, discrete slow oscillation and spindle events were detected using established procedures adapted from previous work^37^. Slow oscillations were identified in the 0.5–1.25 Hz range and sleep spindles in the 12–16 Hz range. Coupled events were defined as spindle events whose midpoint occurred within a detected slow oscillation. Further details regarding spectral analyses, event detection algorithms, and SO–spindle coupling procedures are provided in the Supplementary Methods.

*Script-driven imagery (SDI)* is a validated symptom-provocation procedure used to assess subjective and physiological responses to autobiographical memories^26^. Prior to the adaptation night, participants provided detailed descriptions of two neutral and two negative autobiographical memories. Based on these narratives, individualized 30-s audio scripts were created and recorded in the present tense using a standardized format. Each SDI trial consisted of four consecutive 30-s phases: bas eline, script listening, imagery, and recovery. During the script-listening phase, participants listened to the individualized audio recording describing the autobiographical event. This was followed by an imagery phase, during which participants were instructed to vividly imagine and mentally re-experience the event. After each trial, participants rated emotional arousal, emotional valence, and vividness of the memory on 10-point Likert scales ranging from 1 (“not at all”) to 10 (“extremely”). To minimize carry-over effects, neutral memories were always presented before aversive memories. SDI presentation and questionnaire administration were implemented using PsychoPy^38^. Subjective ratings and heart rate responses were analysed as difference scores between negative and neutral memories. Heart rate was calculated separately for each SDI phase. Consistent with previous SDI studies, analyses focused on responses obtained during the imagery phase, which provides a standardized measure of memory-related emotional reactivity.

#### Psychotherapeutic Intervention

*Imagery Rescripting and Reprocessing Therapy (IRRT)*. In Study 1, participants received a single session of imagery rescripting and reprocessing therapy (IRRT) administered by a trained psychologist according to the protocol described by Schmucker and Köster (2024)^39^. The intervention consisted of three phases. First, participants vividly relived and described the aversive autobiographical memory in imagination (imaginal exposure). Second, they re-entered the memory as their current adult self and were encouraged to intervene in the scene, confront the perpetrator, and modify the course of events until the situation was experienced as resolved (mastery imagery). Third, participants imagined an interaction between their current and younger self, during which the adult self-provided reassurance, protection, and emotional support (self-calming). The intervention aimed to modify maladaptive emotional representations of the memory and promote emotional relief.

#### Odour cueing

Odour cues were delivered using a computer-controlled olfactometer (Ezzence^40^). To establish the cue–memory association, odour presentation was initiated at the beginning of Phase 2 (“Mastery Imagery”) of the IRRT procedure and terminated at the end of Phase 3 (“Self-Calming”). During this period, odours were delivered in 30-ms bursts separated by interstimulus intervals of 30 s. During subsequent sleep, participants were re-exposed either to the same odour that had been paired with IRRT (congruent condition) or to a novel odour not associated with the therapeutic intervention (incongruent condition). Odour presentation alternated between 4-min ON and 4-min OFF intervals. During odour ON periods, odours were administered in 20-ms bursts with an interstimulus interval of 55 s. Odour stimulation was initiated manually during stable N3 sleep and terminated approximately 30 min before scheduled awakening to minimize cue presentation during wakefulness.

*Statistical analyses* were performed using SPSS (IBM Corp.) and MATLAB with the FieldTrip toolbox^41^. Changes in subjective ratings and heart rate were analysed using repeated-measures analyses of variance (ANOVAs) with condition (congruent vs. incongruent odour) and time (T0, T1, T2) as within-subject factors. Differences in sleep architecture between congruent and incongruent nights were assessed using paired-samples *t*-tests.

Cueing-related changes in sleep EEG activity were examined using non-parametric cluster-based permutation statistics implemented in FieldTrip^42^. Analyses were conducted separately for the congruent and incongruent conditions by comparing odour ON and OFF periods across all electrodes. For each oscillatory measure (SWA, spindle activity, REM theta activity, and SO–spindle coupling), paired-samples *t*-tests were first computed at each electrode (sample-level α = 0.05). Spatially adjacent electrodes exceeding this threshold were grouped into clusters, and cluster-level statistics were calculated as the sum of *t*-values within each cluster. Statistical significance was determined using 5,000 permutations, with observed clusters evaluated against the distribution of maximum cluster statistics (cluster-level α = 0.05).

### STUDY 2

*Participants* were eligible if they (1) were between 18 and 60 years of age, (2) met diagnostic criteria for PTSD according to the Clinician-Administered PTSD Scale for DSM-5 (CAPS-5)^29^, (3) had experienced at least one traumatic event before the age of 16 years, and (4) provided informed consent to participate in trauma-focused treatment as part of the study. Exclusion criteria were (1) a lifetime diagnosis of a psychotic or bipolar disorder, (2) intellectual impairment (IQ < 85), (3) current substance use disorder, including benzodiazepine or opioid misuse, (4) acute suicidality, (5) neurological disease (e.g., epilepsy), brain injury, or neurodegenerative disorder (e.g., dementia or Parkinson’s disease), (6) olfactory dysfunction (e.g., due to allergies or post-COVID condition), and (7) prior PTSD treatment exceeding two sessions. Treatment adherence was high, with most participants completing the 12-session protocol, supporting the feasibility of home-based odour cueing as an adjunct to routine outpatient care. Between April 2021 and November 2023, 54 patients were assessed at baseline. Five patients were nonstarters and 49 patients were randomized to conditions. Ten patients dropped out of treatment: three (7.9%) from the congruent condition and seven (18.4%) from the incongruent condition (*χ²* = 1.63, exact *p* = .292). Reasons for dropout included rejection of the treatment program or framework of the study (n = 5), discontinuation of treatment due to nonattendance (n = 2), change in life circumstances (n = 2), and one patient had to be excluded from the analyses due to non-adherence to the use of the odour diffusor (n = 1). For demographic and clinical characteristics of the sample, as well as treatment information, see Table S1. For information on randomization and masking as well as sample size calculation please refer to the Supplementary Methods.

*Primary outcomes* The primary outcome was PTSD symptom severity at the 1-month follow-up assessed with the Clinician-Administered PTSD Scale for DSM-5 (CAPS-5)^29^. The CAPS-5 is a structured clinical interview assessing the frequency and intensity of PTSD symptoms during the preceding month. Total scores range from 0 to 80, with higher scores indicating greater symptom severity. In addition to total scores, symptom cluster scores for intrusion, avoidance, negative alterations in cognition and mood, and hyperarousal were analysed. For secondary psychological outcomes please refer to the Supplementary Methods.

#### Sleep macrostructure and sleep EEG oscillatory activity

Subjective sleep quality was assessed using the Pittsburgh Sleep Quality Index (PSQI^43^). For participants who agreed to laboratory sleep assessments, overnight polysomnography including electroencephalography (EEG) was conducted at baseline and 1-month follow-up. Sleep was recorded using a 7-channel EEG (F4, Fz, C3, C4, Cz, O2, and Oz), together with EOG and EMG, using a SOMNOmedics recording system (SOMNOmedics AG, Randersacker, Germany). Oz served as the online reference during data acquisition. For offline analyses, EEG signals were re-referenced to the linked mastoids (A1 and A2). Sleep architecture and EEG oscillatory activity, including slow-wave activity (SWA), slow oscillations (SO), and fast spindle activity, were analysed using the same procedures as described for Study 1 (see methods Study 1 and Supplementary Methods).

#### Psychotherapeutic intervention: Imagery Rescripting

The intervention consisted of 12 weekly 90-min sessions of imagery rescripting (ImRs). During the first session, patients received psychoeducation regarding the rationale of ImRs, created a hierarchy of traumatic memories, and completed a practice rescripting exercise using a less aversive, non-traumatic memory to familiarize themselves with the procedure. Sessions 2–12 focused on trauma processing using ImRs. Treatment followed the protocol developed by Arntz and Weertman^28,44^ and tested in a clinical trial^45^. During the initial phase of each rescripting exercise, patients were instructed to vividly relive the traumatic memory from the perspective of their younger self while describing the experience in the present tense. Therapists focused on activating the sensory, emotional, cognitive, and interpersonal aspects of the memory, including unmet needs associated with the traumatic experience. Once the memory and associated affect had been sufficiently activated, a helping person entered the imagined scene to intervene, stop the threat or abuse, and provide protection, support, and need fulfillment. During sessions 1–6, the therapist assumed the role of the helping person. During sessions 7–12, patients entered the imagery themselves in their current adult form and performed the rescripting. Therapists were instructed to address the designated index trauma within the first six treatment sessions. All therapists were licensed psychologists, psychotherapists, or psychiatrists who had received formal training in ImRs.

#### Odour cueing

Starting with session 2, patients were exposed to an odour cue delivered via an commercially available odour diffuser during imagery rescripting sessions. Patients subsequently received a home diffuser containing either the same odour that had been presented during therapy (congruent condition) or a different odour not associated with the therapeutic intervention (incongruent condition). Participants were instructed to use the diffuser during sleep for at least two nights following each treatment session to promote reactivation of therapy-related memories during subsequent sleep.

*Statistical analyses* were conducted using SPSS version 29. All tests were two-sided and statistical significance was defined as *p* < 0.05. Analyses were performed on treatment completers. Missing values were not imputed, as generalized linear mixed models (GLMMs) provide valid estimates under assumptions comparable to those underlying multiple imputation approaches. Because several outcome variables showed skewed distributions, continuous outcomes were analysed using GLMMs with either negative binomial or gamma distributions, depending on the characteristics of the data. For variables containing zero values, a constant of 0.01 was added prior to gamma regression analyses. Time was modelled according to the best-fitting trajectory (e.g., linear or logarithmic), and covariance structures were selected based on the lowest corrected Akaike Information Criterion (AICC) among candidate covariance matrices (AR1, ARH1, CS, CSH, ARMA11, and UN).

Time-to-event analyses based on weekly PCL-5 assessments examined remission, response, and recovery as outcomes. Participants who did not reach the respective outcome during treatment were censored. Kaplan–Meier analyses were used to estimate time to remission, response, and recovery, with between-group differences assessed using log-rank tests. Hazard ratios were estimated using Cox proportional hazards regression.

For analyses of sleep EEG activity, slow-wave activity and fast spindle activity were analysed using repeated-measures ANOVAs with the within-subject factors time, electrode, and condition (congruent vs. incongruent). EEG measures were log-transformed prior to analysis to improve normality.

Between group effect sizes were calculated as standardized mean differences (SMD). They were calculated as the difference between transformed estimated mean scores of the treatment groups at each time point, divided by the pooled standard deviation (transformed scale) at each time point, based on a GLMM with gamma or negative binominal regression with only a fixed intercept. Effect sizes were interpreted using conventional criteria, with *d* values of 0.2, 0.5, and 0.8 reflecting small, medium, and large effects, respectively^46^.

## Data availability

The data supporting the findings of this study are available from the corresponding author upon reasonable request. Individual participant data underlying the results reported in this article may be shared with researchers who provide a methodologically sound proposal to IWG. Proposals may be submitted for up to 36 months after publication of this article.

## Statements

### Ethics and Trial Registration

The study was reviewed and approved by the ethics committee of Lübeck University (reference number 19-326) and registered in the German Clinical Trials Register under identifier DRKS00026210.

### Role of funding source

This work was supported by the Swiss National Science Foundation (SNSF; project no. 10001C_179241 to I.W.) and the German Research Foundation (Deutsche Forschungsgemeinschaft, DFG; FOR 5434, WI 4059/2-1, project no. 468645090 to I.W.; TRR 418, project no. 565232769 to I.W.). The funding bodies had no role in the study design, data analysis, data interpretation, or the decision to submit the manuscript for publication.

### Author contributions (ICMJE)

Sabine Groch and Ines Wilhelm designed the study with substantial input from Eva Fassbinder, Anja Schaich, and Judith Amores. Mojgan Ehsanifard, Anja Schaich, Sabine Groch, and Clara Sayk collected the data. Anja Schaich, Mojgan Ehsanifard, Clara Sayk, and Jovana Lehmann-Grube analysed the data with substantial input from Ines Wilhelm and Hong-Viet V Ngo-Dehning. Ines Wilhelm, Anja Schaich, Mathias Kammerer and Sabine Groch interpreted the data with input from Jan-Philipp Klein, Frieder Paulus, Mathias Kammerer and Sören Krach. Anja Schaich and Ines Wilhelm wrote the manuscript with substantial input from all the authors. All authors commented on the manuscript and approved the final version.

### Conflicts of Interest

The authors declare no competing interests.

## Acknowledgments

The authors wish to thank all the patients and therapists who participated in this trial.

## Notes

### Competing Interest Statement

The authors have declared no competing interest.

### Clinical Trial

DRKS00026210

### Author Declarations

The study was reviewed and approved by the ethics committee of Luebeck University (reference number 19-326).

## References

1. American Psychiatric Association,. Diagnostic and Statistical Manual of Mental Disorders (DSM-5®). (>American Psychiatric Pub, 2013).

2. Măirean, C. et al. PTSD symptoms and quality of life after childhood traumatic experiences: A meta-analysis. J. Loss Trauma 29, 377–403 (2024).

3. Scoglio, A. A. J. et al. Social functioning in individuals with post-traumatic stress disorder: A systematic review. Trauma Violence Abuse 23, 356–371 (2022).

4. Baltjes, F., Cook, J. M., van Kordenoordt, M. & Sobczak, S. Psychiatric comorbidities in older adults with posttraumatic stress disorder: A systematic review. Int. J. Geriatr. Psychiatry 38, e5947 (2023).

5. Moder, J. et al. Post-traumatic stress disorder and its somatic comorbidities: A review. J. Educ. Health Sport 80, 59387 (2025).

6. Davis, L. L. et al. The economic burden of posttraumatic stress disorder in the United States from a societal perspective. J. Clin. Psychiatry 83, 21m14116 (2022).

7. Burback, L. et al. Evolving psychotherapeutic approaches for PTSD: Beyond the fear-based model. Psyc.. Clin. Psychopharmacol. 35, S152–S167 (2025).

8. Semmlinger, V. et al. Prevalence and predictors of nonresponse to psychological treatment for PTSD: A meta-analysis. Depress. Anxiety 2024, 9899034 (2024).

9. Varker, T. et al. Dropout from guideline-recommended psychological treatments for posttraumatic stress disorder: A systematic review and meta-analysis. J. Affect. Disord. Rep. 4, 100093 (2021).

10. van der Heijden, A. C., van den Heuvel, O. A., van der Werf, Y. D., Talamini, L. M. & van Marle, H. J. F. Sleep as a window to target traumatic memories. Neurosci. Biobehav. Rev. 140, 104765 (2022).

11. Azza, Y., Wilhelm, I. & Kleim, B. Sleep early after trauma: A target for prevention and early intervention for posttraumatic stress disorder? Eur. Psychol. 25, 239–251 (2020).

12. Phelps, E. A. & Hofmann, S. G. Memory editing from science fiction to clinical practice. Nature 572, 43–50 (2019).

13. Xia, T. & Hu, X. Memory editing during sleep: mechanisms, clinical applications, and technological innovations. Trends Cogn. Sci. 30, 335–349 (2026).

14. Brodt, S., Inostroza, M., Niethard, N. & Born, J. Sleep-A brain-state serving systems memory consolidation. Neuron 111, 1050–1075 (2023).

15. Azza, Y. et al. Sleep’s role in updating aversive autobiographical memories. Transl. Psychiatry 12, 117 (2022).

16. Lutz, N. D., Harkotte, M. & Born, J. Sleep’s contribution to memory formation. Physiol. Rev. 106, 363–483 (2026).

17. Born, J. & Wilhelm, I. System consolidation of memory during sleep. Psychol. Res. 76, 192–203 (2012).

18. Rasch, B., Büchel, C., Gais, S. & Born, J. Odor cues during slow-wave sleep prompt declarative memory consolidation. Science 315, 1426–1429 (2007).

19. Groch, S. et al. Targeted reactivation during sleep differentially affects negative memories in socially anxious and healthy children and adolescents. J. Neurosci. 37, 2425–2434 (2017).

20. Hu, X., Cheng, L. Y., Chiu, M. H. & Paller, K. A. Promoting memory consolidation during sleep: A meta-analysis of targeted memory reactivation. Psychol. Bull. 146, 218–244 (2020).

21. Groch, S., Schreiner, T., Rasch, B., Huber, R. & Wilhelm, I. Prior knowledge is essential for the beneficial effect of targeted memory reactivation during sleep. Sci. Rep. 7, 39763 (2017).

22. Rihm, J. S., Sollberger, S. B., Soravia, L. M. & Rasch, B. Re-presentation of olfactory exposure therapy success cues during non-rapid eye movement sleep did not increase therapy outcome but increased sleep spindles. Front. Hum. Neurosci. 10, 340 (2016).

23. Borghese, F. et al. Targeted memory reactivation during REM sleep in patients with social anxiety disorder. Front. Psychiatry 13, 904704 (2022).

24. Recher, D. et al. Targeted memory reactivation during sleep improves emotional memory modulation following imagery rescripting. Transl. Psychiatry 14, 490 (2024).

25. van der Heijden, A. C., van der Werf, Y. D., van den Heuvel, O. A., Talamini, L. M. & van Marle, H. J. F. Targeted memory reactivation to augment treatment in post-traumatic stress disorder. Curr. Biol. 34, 3735–3746.e5 (2024).

26. Pitman, R. K., Orr, S. P., Forgue, D. F., B., de J. J. & Claiborn, J. M. Psychophysiologic assessment of posttraumatic stress disorder imagery in Vietnam combat veterans. Archives of general psychiatry 44, 970–975 (1987).

27. Smucker, M. R. Imagery rescripting and reprocessing therapy. in *Encyclopedia of Cognitive Behavior Therapy* 226–229 (Springer-Verlag, New York, 2006).

28. Arntz, A. Imagery Rescripting: an update of the treatment protocol. Behav. Res. Ther. 195, 104913 (2025).

29. Weathers, F. W. et al. The Clinician-Administered PTSD Scale for DSM-5 (CAPS-5): Development and initial psychometric evaluation in military veterans. Psychol. Assess. 30, 383–395 (2018).

30. Blevins, C. A., Weathers, F. W., Davis, M. T., Witte, T. K. & Domino, J. L. The Posttraumatic Stress Disorder Checklist for DSM-5 (PCL-5): Development and initial psychometric evaluation: Posttraumatic stress disorder checklist for DSM-5. J. Trauma. Stress 28, 489–498 (2015).

31. Lane, R. D., Ryan, L., Nadel, L. & Greenberg, L. Memory reconsolidation, emotional arousal, and the process of change in psychotherapy: New insights from brain science. Behav. Brain Sci. 38, e1 (2015).

32. Klinzing, J. G., Niethard, N. & Born, J. Mechanisms of systems memory consolidation during sleep. Nat. Neurosci. 22, 1598–1610 (2019).

33. Schreiner, T., Petzka, M., Staudigl, T. & Staresina, B. P. Endogenous memory reactivation during sleep in humans is clocked by slow oscillation-spindle complexes. Nat. Commun. 12, 3112 (2021).

34. Helfrich, R. F., Mander, B. A., Jagust, W. J., Knight, R. T. & Walker, M. P. Old brains come uncoupled in sleep: Slow wave-spindle synchrony, brain atrophy, and forgetting. Neuron 97, 221–230.e4 (2018).

35. Ngo, H.-V. V., Martinetz, T., Born, J. & Mölle, M. Auditory closed-loop stimulation of the sleep slow oscillation enhances memory. Neuron 78, 545–553 (2013).

36. Diekelmann, S., Wilhelm, I. & Born, J. The whats and whens of sleep-dependent memory consolidation. Sleep medicine reviews 13, 309–321 (2009).

37. Klinzing, J. G. et al. Auditory stimulation during sleep suppresses spike activity in benign epilepsy with centrotemporal spikes. Cell Rep. Med. 2, 100432 (2021).

38. Peirce, J. et al. PsychoPy2: Experiments in behavior made easy. Behav. Res. Methods 51, 195–203 (2019).

39. Schmucker, M. & Köster, R. Praxishandbuch IRRT (Leben Lernen, Bd. 351): Imagery Rescripting & Reprocessing Therapy bei Traumafolgestörungen, Angst, Depression und Trauer. (2025).

40. Amores, J., Dotan, M. & Maes, P. Development and study of Ezzence: A modular scent wearable to improve wellbeing in home sleep environments. Front. Psychol. 13, 791768 (2022).

41. Oostenveld, R., Fries, P., Maris, E. & Schoffelen, J.-M. FieldTrip: Open source software for advanced analysis of MEG, EEG, and invasive electrophysiological data. Comput. Intell. Neurosci. 2011, 156869 (2011).

42. Maris, E. & Oostenveld, R. Nonparametric statistical testing of EEG- and MEG-data. J. Neurosci. Methods 164, 177–190 (2007).

43. Smyth, C. The Pittsburgh sleep quality index (PSQI). J. Gerontol. Nurs. 25, 10–11 (1999).

44. Arntz, A. & Weertman, A. Treatment of childhood memories: Theory and practice. Behav. Res. Ther. 37, 715–740 (1999).

45. Boterhoven de Haan, K. L., et al. Imagery rescripting and eye movement desensitisation and reprocessing as treatment for adults with post-traumatic stress disorder from childhood trauma: randomised clinical trial. Br. J. Psychiatry 217, 609–615 (2020).

46. Cohen, J. Statistical Power Analysis for the Behavioral Sciences. (Academic press, San Diego, CA, 2013).

