## Supplement for "Odour cueing during sleep augments trauma-focused psychotherapy"

### Supplementary Results

**Table S1.** Test statistics of the ANOVA of the SDI outcomes in study 1

| | | ANOVA Analyses |
|---|---|---|
| SDI: arousal | Time | $F(2, 25) = 35.97, p < .001$ |
| | Condition | $F(1, 26) = 0.18, p = .677$ |
| | Time*Condition | $F(2, 25) = 0.53, p = .596$ |
| SDI: negative emotional valence | Time | $F(2, 23) = 32.97, p < .001$ |
| | Condition | $F(1,24) = 0.00, p = .967$ |
| | Time*Condition | $F(2, 23) = 0.37, p = .696$ |
| SDI: vividness | Time | $F(2, 25) = 0.45, p = .646$ |
| | Condition | $F(1, 26) = 0.39, p = .537$ |
| | Time*Condition | $F(2, 25) = 2.06, p = .149$ |
| HR: change score emotional - neutral | Time | $F(2, 21) = 1.44, p = .259$ |
| | Condition | $F(1, 22) = 0.25, p = .624$ |
| | Time*Condition | $F(2, 21) = 0.83, p = .448$ |

*Abbreviations:* ANOVA, analysis of variance; SDI, Script Driven Imagery; HR, mean heart rate

**Table S2.** Descriptive statistics of the SDI outcomes in study 1

| Outcome | Assessment | M (SD) | |
|---|---|---|---|
| | | Congruent | Incongruent |
| SDI: arousal change score emotional - neutral | pre | 3.88 (1.63) | 4.29 (1.58) |
| | post1 | 2.37 (1.24) | 2.29 (1.58) |
| | post2 | 2.07 (1.52) | 2.11 (1.36) |
| SDI: negative emotional valence change score emotional - neutral | pre | 4.88 (1.69) | 4.56 (2.16) |
| | post1 | 2.52 (1.69) | 2.72 (1.86) |
| | post2 | 2.08 (1.99) | 2.24 (1.56) |
| SDI: vividness change score emotional - neutral | pre | -0.71 (1.58) | -0.89 (1.64) |
| | post1 | -0.82 (1.51) | -0.32 (2.24) |
| | post2 | -0.78 (1.83) | -0.71 (2.17) |
| HR: change score emotional - neutral | pre | 0.34 (3.12) | -0.18 (3.83) |
| | post1 | 0.04 (3.63) | 0.82 (4.63) |
| | post2 | -1.06 (3.05) | -0.39 (3.14) |
| HR: emotional memory | pre | 70.69 (11.04) | 71.06 (10.05) |
| | post1 | 67.16 (8.89) | 68.48 (8.47) |
| | post2 | 73.95 (9.48) | 72.28 (11.42) |
| HR: neutral memory | pre | 70.35 (10.6) | 71.24 (10.67) |
| | post1 | 67.12 (8.99) | 67.67 (8.97) |
| | post2 | 75.01 (9.22) | 72.67 (11.44) |

*Abbreviations:* SDI, Script Driven Imagery; HR, mean heart rate

**Figure S1**. Slow Wave Activity in congruent and incongruent cueing condition

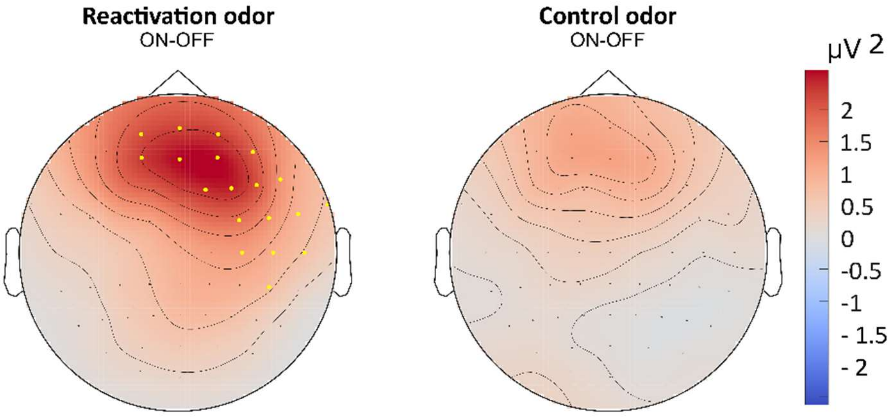

Topoplots depict differences between odour ON and OFF periods for the congruent condition (left) and incongruent condition (right). Significant electrode clusters ($p < 0.05$) are highlighted in yellow.

**Table S3.** Test statistics of the paired sample t-tests for sleep architecture in study 1

| | Condition | | | | | |
|---|---|---|---|---|---|---|
| | Congruent | | Incongruent | | | |
| | M | *SD* | M | *SD* | *p* | *Hedge's g* |
| Sleep latency (stage 2), min | 12.41 | 16.56 | 14.74 | 11.93 | .371 | -.192 |
| Sleep efficiency, % | 93.57 | 0.05 | 93.28 | 0.05 | .756 | .066 |
| Total sleep duration, min | 422.95 | 35.35 | 428.26 | 34.08 | .427 | -.170 |
| WASO duration, min | 14.92 | 12.88 | 13.95 | 15.44 | .782 | .059 |
| Slow wave sleep (N3), % | 14.09 | 5.45 | 15.02 | 6.16 | .363 | -.196 |
| REM sleep, % | 21.93 | 3.78 | 21.77 | 5.06 | .906 | .025 |
| MA Slow Wave Sleep (N3) | 0.62 | 0.72 | 1.21 | 1.29 | .004 | -.689 |
| MA REM Sleep | 6.14 | 3.35 | 6.69 | 4.03 | .359 | -.197 |
| MA Total | 23.57 | 8.72 | 23.42 | 10.19 | .913 | .023 |

*Abbreviations:* WASO, wake after sleep onset; REM, rapid eye movement; MA, movement arousal

**Table S4**. Correlations between significant electrode clusters and SDI measures in study 1

| | Arousal post1 | Arousal post2 | Valence post1 | Valence post2 | Vividness post1 | Vividness post2 | HR post1 | HR post2 |
|---|---|---|---|---|---|---|---|---|
| SO-Spindle coupling (congruent) | -.472* | -.170 | -.376 | -.155 | -.282 | -.280 | -.180 | .253 |
| SO-Spindle coupling (incongruent) | -.038 | .281 | -.075 | .238 | -.222 | .129 | -.104 | -.147 |
| SO (congruent) | .094 | -.085 | .060 | -.186 | -.158 | -.230 | -.099 | -.081 |
| SO (incongruent) | .147 | .161 | .285 | .326 | .150 | .277 | .107 | .439 |
| SWA (congruent) | .109 | -.091 | .041 | -.281 | -.067 | -.160 | -.125 | -.056 |
| SWA (incongruent) | -.114 | -.043 | .030 | .032 | .163 | .104 | .043 | -.093 |

Abbreviations: Script-driven imagery, SDI; Heart rate, HR; Slow Oscillations, SO; Slow Wave Activity, SWA; * $p < 0.05$

**Figure S2.** CONSORT diagram of participant flow in study 2.

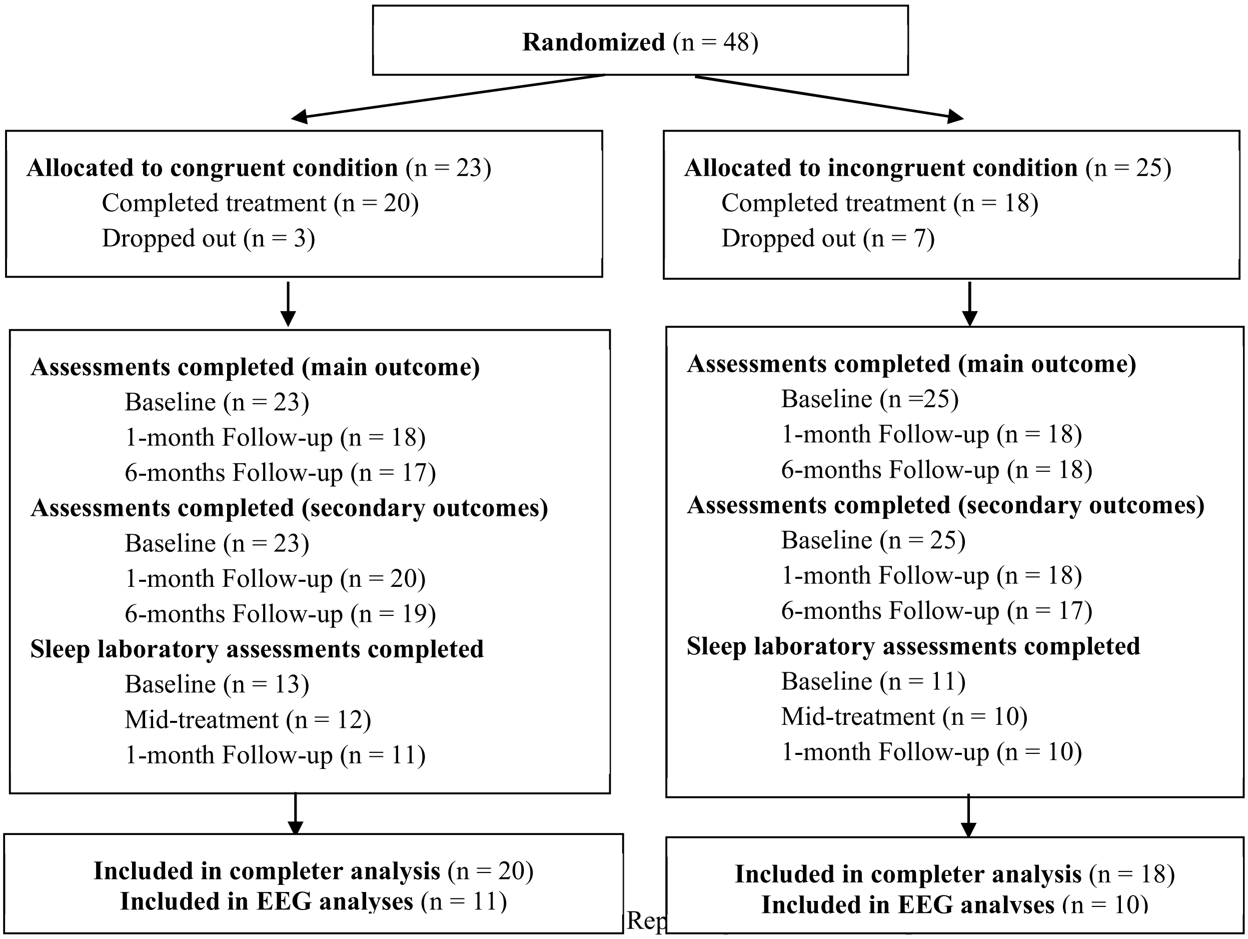

EEG: electroencephalography

**Table S5.** Sample characteristics of treatment completers and treatment information of study 2

| | Total ($N$ = 38) | Congruent ($n$ = 20) | Incongruent ($n$ = 18) | Congruent vs. incongruent |
|---|---|---|---|---|
| Age, *M (SD)* | 38.00 (10.96) | 37.50 (10.81) | 38.56 (11.43) | $U$ = 189.50 |
| Gender, *n (%)* | | | | $\chi^2 = 0.26$ |
| female | 35 (92.1) | 18 (90.0) | 17 (94.4) | |
| male | 3 (7.9) | 2 (10.0) | 1 (5.6) | |
| Family status, *n (%)* | | | | $\chi^2 = 0.00$ |
| Single | 17 (44.7) | 9 (45.0) | 8 (44.4) | |
| In relationship | 21 (55.3) | 11 (55.0) | 10 (55.6) | |
| Education level, *n (%)* | | | | $\chi^2 = 2.72$ |
| Lower secondary education | 23 (60.5) | 13 (65.0) | 10 (55.6) | |
| Upper secondary education | 7 (18.4) | 2 (10.0) | 5 (28.8) | |
| Tertiary education | 7 (18.4) | 4 (20.0) | 3 (16.7) | |
| Other | 1 (2.6) | 1 (5.0) | 0 (0.0) | |
| Work status, *n (%)* | | | | $\chi^2 = 6.70$ |
| Working | 16 (42.0) | 10 (50.0) | 6 (33.3) | |
| Unpaid work | 1 (2.6) | 0 (0.0) | 1 (5.6) | |
| Student | 4 (10.5) | 1 (5.0) | 3 (16.7) | |
| Homemaker | 2 (5.3) | 2 (10.0) | 0 (0.0) | |
| Unemployed/unable to work | 7 (18.4) | 2 (10.0) | 5 (27.8) | |
| Other | 8 (21.1) | 5 (25.0) | 3 (16.7) | |
| Psychotropic medication at baseline, *n (%)* | | | | $\chi^2 = 3.70$ |
| Psychotropic medication | 23 (60.5) | 15 (75.0) | 8 (44.4) | |
| No psychotropic medication | 15 (42.1) | 5 (25.0) | 10 (55.6) | |
| Duration of therapy, *M (SD)* | | 11.45 | | |
| Number of weeks | | | | |
| Number of ImRs sessions | | | | |
| Treatments with a 13th session[1] | 6 (15.8) | 2 (10.0) | 4 (22.2) | $\chi^2 = 1.06$ |
| Early successes, *n (%)* | 6 (15.8) | 6 (30.0) | 0 (0.0) | $\chi^2 = 6.41$* |
| Blinding of participants[2], *n (%)* | | | | |
| Guessed aim of the study correctly | 2 (5.3) | 2 (10.0) | 0 (0.0) | $\chi^2 = 2.00$ |
| Guessed assigned condition correctly | 1 (2.6) | 1 (5.0) | 0 (0.0) | $\chi^2 = 0.97$ |
| Guessed that odours were the same | 23 (60.5) | 12 (60.0) | 11 (61.1) | |
| Guessed that odours were different | 6 (15.8) | 4 (20.0) | 2 (11.1) | $\chi^2 = 2.68$ |
| Doesn't know if odours were the same | 2 (5.3) | 0 (0.0) | 2 (11.1) | |
| Missings | 7 (18.4) | 4 (20.0) | 3 (16.7) | |
| Sleep laboratory assessments | Total ($N$ = 21) | Congruent ($n$ = 11) | Incongruent ($n$ = 10) | Congruent vs. incongruent |
| Age, *M (SD)* | 36.82 (10.67) | 34.42 (9.38) | 39.70 (11.89) | $U$ = 42.50 |
| Gender, *n (%)* | | | | $\chi^2 = 0.21$ |
| female | 19 (92.1) | 10 (83.3) | 9 (90) | |
| male | 3 (7.9) | 2 (16.7) | 1 (10) | |

[1]If Imagery Rescripting could not be conducted in one of the 12 sessions for any reason, a 13th session was allowed, [2]assessed by a telephone interview after completion of the study, * p < .05, ** p < .01, *** p < .001

**Table S6:** Test statistics of the General Linear Mixed Model analyses of primary and secondary outcomes in study 2.

| | | GLMM Analyses |
|---|---|---|
| CAPS: Total score | Time | $F(2, 32) = 26.06, p < .001$ |
| | Condition | $F(1, 32) = 2.04, p = .163$ |
| | Time*Condition | $F(2, 32) = 2.77, p = .077$ |
| CAPS: Intrusions | Time | $F(2, 34) = 17.34, p <.001$ |
| | Condition | $F(1,35) = 4.68, p = .038$ |
| | Time*Condition | $F(2, 34) = 3.40, p = .045$ |
| CAPS: Avoidance | Time | $F(2, 34) = 14.49, p <.001$ |
| | Condition | $F(1, 34) = 0.22, p = .646$ |
| | Time*Condition | $F(2, 34) = 1.19, p = .316$ |
| CAPS: Emotions and cognitions | Time | $F(2, 41) = 28.37, p <.001$ |
| | Condition | $F(1, 34) = 1.84, p = .184$ |
| | Time*Condition | $F(2, 41) = 2.52, p = .093$ |
| CAPS: Arousal and reactivity | Time | $F(2, 33) = 12.03, p <.001$ |
| | Condition | $F(1, 32) = 1.70, p = .201$ |
| | Time*Condition | $F(2, 33) = 2.84, p = .073$ |
| CAPS: Dissociation | Time | $F(2, 33) = 3.06, p = .060$ |
| | Condition | $F(1, 33) = 0.00, p = .954$ |
| | Time*Condition | $F(2, 33) = 0.25, p = .779$ |
| PCL (index trauma): Total score | Time | $F(15, 466) = 13.08, p < .001$ |
| | Condition | $F(1, 37) = 4.90, p = .033$ |
| | Time*Condition | $F(15, 466) = 1.88, p = .024$ |
| PCL (index trauma): Re-experiencing | Time | $F(15, 322) = 3.58, p < .001$ |
| | Condition | $F(1, 41) = 6.38, p = .016$ |
| | Time*Condition | $F(15, 322) = 2.43, p = .002$ |
| PCL (index trauma): Avoidance | Time | $F(15, 320) = 4.17, p < .001$ |
| | Condition | $F(1, 42) = 7.12, p = .011$ |
| | Time*Condition | $F(15, 320) = 1.50, p = .102$ |
| PCL (index trauma): Negative alterations in cognitions and mood | Time | $F(15, 125) = 2.59, p = .002$ |
| | Condition | $F(1, 52) = 7.95, p = .007$ |
| | Time*Condition | $F(15, 125) = 1.91, p = .028$ |
| PCL (index trauma): Hyperarousal | Time | $F(15, 295) = 3.09, p < .001$ |
| | Condition | $F(1, 43) = 1.87, p = .179$ |
| | Time*Condition | $F(15, 295) = 0.83, p = .643$ |
| PTCI | Time | $F(15, 157) = 1.79, p = .040$ |
| | Condition | $F(1, 48) = 2.07, p = .157$ |
| | Time*Condition | $F(15, 157) = 1.07, p = .391$ |
| BDI-II | Time | $F(2, 45) = 14.69, p < .001$ |
| | Condition | $F(1, 36) = 0.41, p = .526$ |
| | Time*Condition | $F(2, 45) = 0.96, p = .391$ |
| WHODAS 2.0 | Time | $F(2, 36) = 6.13, p =.005$ |
| | Condition | F(1, 36) = 4.65, $p = .038$ |
| | Time*Condition | $F(2, 36) = 1.82, p = .177$ |
| GSE | Time | $F(2, 67) = 7.26, p =.001$ |
| | Condition | $F(1, 36) = 4.37, p = .044$ |
| | Time*Condition | $F(2, 67) = 0.40, p = .669$ |
| BAI | Time | $F(2, 70) = 7.10, p = .002$ |
| | Condition | $F(1, 36) = 1.75, p = .194$ |
| | Time*Condition | $F(2, 70) = 0.09, p = .915$ |
| PSQI | Time | $F(2, 69) = 12.79, p < .001$ |
| | Condition | $F(1, 40) = 0.14, p = .709$ |
| | Time*Condition | $F(2, 69) = 0.39$, p = .678 |
| Sleep latency (stage 2), min | Time | $F(1, 16) = 0.28, p = .606$ |
| | Condition | $F(1, 17) = 0.29, p = .599$ |
| | Time*Condition | $F(1, 16) = 0.46, p = .509$ |
| Sleep efficiency, % | Time | $F(1, 17) = 0.35, p = .564$ |

| | | |
|---|---|---|
| | Condition | $F(1, 17) = 2.43$, $p = .138$ |
| | Time*Condition | $F(1, 17) = 0.04$, $p = .854$ |
| Total sleep duration, min | Time | $F(1, 18) = 0.74$, $p = .403$ |
| | Condition | $F(1, 17) = 0.83$, $p = .376$ |
| | Time*Condition | $F(1, 18) = 0.24$, $p = .629$ |
| WASO duration, % | Time | $F(1, 16) = 0.34$, $p = .566$ |
| | Condition | $F(1, 17) =0.69$, $p = .417$ |
| | Time*Condition | $F(1, 16) = 0.15$, $p = .904$ |
| Non-rem sleep (N1), % | Time | $F(1, 18) = 2.95$, $p = .103$ |
| | Condition | $F(1, 20) = 0.70$, $p = .413$ |
| | Time*Condition | $F(2, 18) = 0.32$, $p = .579$ |
| Non-rem sleep (N2), % | Time | $F(1, 19) = 1.04$, $p = .313$ |
| | Condition | $F(1, 19) = 0.76$, $p = .393$ |
| | Time*Condition | $F(2, 19) = 0.10$, $p = .754$ |
| Slow wave sleep (N3), % | Time | $F(1, 19) = 0.84$, $p = .372$ |
| | Condition | $F(1, 20) = 2.49$, $p = .130$ |
| | Time*Condition | $F(2, 19) = 4.73$, $p = .042$ |
| REM sleep, % | Time | $F(1, 16) = 0.35$, $p = .562$ |
| | Condition | $F(1, 16) =0.79$, $p = .388$ |
| | Time*Condition | $F(2, 16) = 0.49$, $p = .496$ |
| REM density | Time | $F(1, 17) = 3.10$, $p = .097$ |
| | Condition | $F(1, 18) =0.26$, $p = .616$ |
| | Time*Condition | $F(2, 17) = 2.23$, $p = .154$ |
| Movement arousal | Time | $F(1, 19) = 1.82$, $p = .194$ |
| | Condition | $F(1, 20) =0.27$, $p = .611$ |
| | Time*Condition | $F(2, 19) = 0.13$, $p = .725$ |

*Abbreviations:* CAPS-5, Clinician-Administered PTSD Scale for DSM-5; PCL-5, Posttraumatic Stress Disorder Checklist, PTCI, Posttraumatic Cognitions Inventory; BDI-II, Becks Depression Inventory; BAI, Becks Anxiety Inventory; ‘WHODAS 2.0, World Health Organization Disability Assessment Schedule 2.0; GSE, General Self-Efficacy Scale; PSQI, Pittsburgh Sleep Quality Index; WASO, wake after sleep onset, REM, rapid eye movement

**Table S7**. Estimated means and effect sizes of changes from baseline to follow-up for secondary outcomes in study 2

| Outcome | Assessment | M (95% CI) | | SMD [95% CI][1] |
|---|---|---|---|---|
| | | Congruent | Incongruent | |
| CAPS (all traumas) Total score | Baseline | 39.00 [34.70; 43.83] | 40.39 [35.72; 45.67] | |
| | 1-month follow-up | 19.70 [14.85; 26.12] | 28.61 [21.62; 37.86] | 0.38 [-0.29; 1.03] |
| | 6-month follow-up | 19.78 [14.81; 26.43] | 24.50 [18.38; 32.66] | 0.22 [-0.45; 0.88] |
| CAPS Intrusions | Baseline | 10.10 [8.84; 11.55] | 10.50 [9.14; 12.07] | |
| | 1-month follow-up | 4.52 [3.37; 6.06] | 7.61 [5.96; 9.71] | 0.51 [-0.17; 1.16] |
| | 6-month follow-up | 5.38 [3.84; 6.98] | 6.67 [5.10; 8.72] | 0.26 [-0.41; 0.92] |
| CAPS Avoidance | Baseline | 4.75 [3.97; 5.69] | 5.11 [4.24; 6.17] | |
| | 1-month follow-up | 2.22 [1.31; 3.78] | 2.11 [1.22; 3.65] | -0.06 [-0.71; 0.60] |
| | 6-month follow-up | 1.78 [0.97; 3.29] | 2.56 [1.45; 4.50] | 0.43 [-0.25; 1.09] |
| CAPS Emotions and cognitions | Baseline | 14.45 [12.23; 17.07] | 14.89 [12.50; 17.74] | |
| | 1-month follow-up | 7.10 [5.15; 9.78] | 10.83 [7.98; 14.72] | 0.41 [-0.26; 1.06] |
| | 6-month follow-up | 6.83 [4.79; 9.75] | 8.56 [6.07; 12.07] | 0.24 [-0.43; 0.90] |
| CAPS Arousal and reactivity | Baseline | 9.70 [8.16; 11.54] | 9.89 [8.25; 11.86] | |
| | 1-month follow-up | 5.54 [4.12; 7.44] | 8.06 [6.15; 10.55] | 0.38 [-0.29; 1.03] |
| | 6-month follow-up | 5.59 [4.25; 7.51] | 6.72 [5.09; 8.89] | 0.18 [-0.49; 0.84] |
| CAPS Dissociation | Baseline | 1.55 [0.95; 2.54] | 1.61 [0.96; 2.70] | |
| | 1-month follow-up | 1.08 [0.55; 2.12] | 1.17 [0.60; 2.29] | 0.09 [-0.57; 0.74] |
| | 6-month follow-up | 1.07 [0.53; 2.70] | 0.90 [0.42; 1.88] | -0.19 [-0.85; 0.48] |
| PCL (index trauma) Total score | Baseline | 45.65 [36.00; 57.88] | 47.33 [36,86; 60,78] | |
| | Week 1 | 43.30 [34.05; 55.07] | 48.22 [37.56; 61.91] | |
| | Week 2 | 38.35 [30.20; 48.70] | 45.54 [35.35; 58.67] | 0.14 [-0.51; 0.78] |
| | Week 3 | 34.55 [27.03; 44.15] | 46.65 [35.94; 59.62] | 0.21 [-0.46; 0.87] |
| | Week 4 | 35.12 [27.46; 44.75] | 46.65 [36.10; 60.28] | 0.27 [-0.40: 0.94] |
| | Week 5 | 31.20 [24.38; 39.92] | 41.39 [32.09; 53.36] | 0.27 [-0.40; 0.93] |
| | Week 6 | 31.01 [24.23; 39.68] | 43.11 [33.55; 55.40] | 0.32 [-0.35; 0.97] |
| | Week 7 | 29.56 [23.08; 37.85] | 42.94 [33.42; 55.19] | 0.40 [-0.27; 1.05] |
| | Week 8 | 27.33 [21.33; 35.03] | 40.32 [31.27; 52.00] | 0.43 [-0.25; 1.09] |
| | Week 9 | 20.43 [15.76; 26.48] | 43.06 [33.51; 55.33] | 0.88 [0.16; 1.56] |
| | Week 10 | 24.55 [19.12; 31.51] | 38.01 [29.45; 49.04] | 0.54 [-0.15; 1.20] |
| | Week 11 | 25.60 [19.75; 33.19] | 32.55 [25.18; 42.08] | 0.24 [-0.46; 0.93] |
| | Week 12 | 26.02 [19.99; 33.87] | 35.15 [27.03; 45.71] | 0.33 [-0.41; 1.05] |
| | (Week 13) | 30.14 [17.83; 50.96] | 37.70 [25.46; 55.80] | 0.19 [-1.55; 1.85] |
| | 1-month follow-up | 19.95 [15.57; 25.56] | 29.72 [23.04; 38.35] | 0.44 [-0.22; 1.07] |
| | 6-month follow-up | 18.97 [14.75; 24.38] | 27.56 [21.28; 35.70] | 0.43 [-0.25; 1.08] |
| PCL Re-experiencing | Baseline | 10.40 [7.80; 13.88] | 10.78 [7.96; 14.59] | |
| | Week 1 | 9.20 [6.87; 12.33] | 11.33 [8.39; 15.31] | |
| | Week 2 | 8.00 [5.94; 10.78] | 10.22 [7.54; 13.86] | 0.24 [-0.40; 0.88] |
| | Week 3 | 6.45 [4.74; 8.77] | 11.33 [8.39; 15.31] | 0.55 [-0.11; 1.19] |
| | Week 4 | 8.65 [6.44; 11.62] | 11.72 [8.68; 15.82] | 0.30 [-0.34; 0.94] |
| | Week 5 | 7.69 [5.68; 10.40] | 10.28 [7.58; 13.93] | 0.28 [-0.37; 0.92] |
| | Week 6 | 7.30 [5.40; 9.87] | 11.00 [8.13; 14.88] | 0.42 [-0.24; 1.05] |
| | Week 7 | 8.13 [6.02;10.97] | 10.78 [7.96; 14.59] | 0.30 [-0.35; 0.94] |
| | Week 8 | 4.19 [2.98; 5.88] | 12.00 [8.90; 16.19] | 1.12 [0.40; 1.80] |
| | Week 9 | 6.00 [4.38; 8.21] | 9.78 [7.20; 13.28] | 0.53 [-0.14; 1.17] |
| | Week 10 | 5.39 [3.91; 7.43] | 9.89 [7.28; 13.42] | 0.64 [-0.03; 1.29] |
| | Week 11 | 5.71 [4.13; 7.90] | 7.94 [5.80; 10.88] | 0.33 [-0.34; 0.99] |
| | Week 12 | 5.76 [4.10; 8.07] | 8.48 [6.19; 11.62] | 0.48 [-0.25; 1.18] |
| | (Week 13) | 6.98 [3.72; 13.10] | 10.70 [6.79; 16.86] | 0.35 [-1.42; 1.99] |
| | 1-month follow-up | 4.45 [3.20; 6.18] | 6.78 [4.91; 9.35] | 0.44 [-0.21; 1.07] |
| | 6-month follow-up | 3.59 [2.53; 5.08] | 6.84 [4.94; 9.47] | 0.64 [-0.04; 1.30] |

| | | | | |
|---|---|---|---|---|
| PCL Avoidance | Baseline | 6.00 [4.71;7.64] | 5.78 [4.46; 7.48] | |
| | Week 1 | 4.95 [3.83; 6.40] | 5.83 [4.51; 7.55] | |
| | Week 2 | 4.25 [3.24;5.58] | 5.67 [4.37; 7.35] | 0.34 [-0.31; 0.97] |
| | Week 3 | 3.85 [2.91; 5.10] | 5.67 [4.37; 7.35] | 0.44 [-0.21; 1.07] |
| | Week 4 | 3.75 [2.82; 4.98] | 5.56 [4.28; 7.22] | 0.39 [-0.26; 1.03] |
| | Week 5 | 3.05 [2.24; 4.14] | 5.22 [4.00; 6.82] | 0.53 [-0.13; 1.16] |
| | Week 6 | 3.20 [2.37; 4.32] | 5.06 [3.86; 6.62] | 0.35 [-0.30; 0.98] |
| | Week 7 | 2.90 [2.12; 3.96] | 4.78 [3.63; 6.29] | 0.51 [-0.15; 1.14] |
| | Week 8 | 2.52 [1.81; 3.52] | 4.35 [3.27; 5.80] | 0.56 [-0.12; 1.21] |
| | Week 9 | 2.12 [1.47; 3.04] | 4.61 [3.49; 6.09] | 0.77 [0.08; 1.43] |
| | Week 10 | 2.17 [1.52; 3.10] | 4.22 [3.17; 5.62] | 0.69 [0.01; 1.33] |
| | Week 11 | 2.82 [2.03; 3.91] | 3.61 [2.67; 4.89] | 0.26 [-0.41; 0.92] |
| | Week 12 | 2.80 [1.99; 3.95] | 3.82 [2.81; 5.18] | 0.33 [-0.40; 1.05] |
| | (Week 13) | 3.10 [1.62; 5.93] | 3.61 [2.27; 5.75] | 0.14 [-1.59; 1.81] |
| | 1-month follow-up | 1.75 [1.19; 2.57] | 3.17 [2.30; 4.35] | 0.62 [-0.05; 1.25] |
| | 6-month follow-up | 1.58 [1.05; 2.37] | 3.19 [2.31; 4.40] | 0.67 [-0.01; 1.33] |
| PCL Negative alterations in cognitions and mood | Baseline | 16.05 [13.60; 18.94] | 17.39 [14.63; 20.67] | |
| | Week 1 | 15.86 [13.09; 19.21] | 17.83 [14.62; 21.75] | |
| | Week 2 | 14.30 [12.14; 16.84] | 17.11 [14.45; 20.26] | 0.17 [-0.48; 0.80] |
| | Week 3 | 12.35 [10.35; 14.73] | 17.28 [14.44; 20.68] | 0.29 [-0.36; 0.94] |
| | Week 4 | 12.70 [9.79; 16.48] | 17.11 [13.13; 22.30] | 0.29 [-0.35; 0.93] |
| | Week 5 | 11.45 [8.85; 14.81] | 15.01 [11.54; 19.52] | 0.27 [-0.39; 0.91] |
| | Week 6 | 10.75 [8.55; 13.52] | 15.67 [12.47; 19.69] | 0.37 [-0.29; 1.01] |
| | Week 7 | 10.01 [7.92; 12.67] | 15.50 [12.31; 19.52] | 0.44 [-0.23; 1.10] |
| | Week 8 | 9.35 [7.12; 12.28] | 14.33 [10.95; 18.75] | 0.44 [-0.22; 1.08] |
| | Week 9 | 7.58 [5.90; 9.74] | 15.17 [11.99; 19.18] | 0.72 [0.04;1.36] |
| | Week 10 | 9.10 [7.12; 11.63] | 13.78 [10.85; 17.50] | 0.44 [-0.24; 1.09] |
| | Week 11 | 9.78 [7.56; 12.66] | 11.89 [9.22; 15.33] | 0.20 [-0.48; 0.87] |
| | Week 12 | 8.76 [6.38; 12.03] | 12.52 [9.28; 16.87] | 0.35 [-0.37; 1.06] |
| | (Week 13) | 9.79 [5.70; 16.79] | 13.71 [8.99; 20.89] | 0.29 [-1.47; 1.93] |
| | 1-month follow-up | 6.85 [4.73; 9.92] | 11.33 [7.93; 16.21] | 0.51 [-0.15; 1.14] |
| | 6-month follow-up | 7.04 [4.90; 10.10] | 9.94 [6.95; 14.23] | 0.38 [-0.29; 1.03] |
| PCL Hyperarousal | Baseline | 13.20 [10.38; 16.79] | 13.39 [10.40; 17.24] | |
| | Week 1 | 12.51 [9.81; 15.96] | 13.22 [10.26; 17.03] | |
| | Week 2 | 11.80 [9.25; 15.06] | 12.28 [9.49; 15.88] | |
| | Week 3 | 10.71 [8.35; 13.75] | 11.39 [8.80; 14.75] | 0.05 [-0.60; 0.71] |
| | Week 4 | 9.60 [7.45; 12.35] | 12.35 [9.54;15.99] | 0.24 [-0.43; 0.90] |
| | Week 5 | 9.37 [7.27; 12.07] | 11.50 [8.89; 14.88] | 0.20 [-0.45;0.85] |
| | Week 6 | 9.47 [7.35; 12.20] | 11.39 [9.20; 15.37] | 0.18 [-0.47;0.82] |
| | Week 7 | 8.72 [6.75; 11.27] | 11.89 [9.20; 15.37] | 0.32 [-0.34;0.96] |
| | Week 8 | 9.00 [6.97; 11.63] | 11.11 [8.57; 14.40] | 0.22 [-0.44;0.87] |
| | Week 9 | 7.56 [5.79; 9.86] | 11.28 [8.71; 14.61] | 0.43 [-0.25; 1.09] |
| | Week 10 | 7.86 [6.05; 10.21] | 10.59 [8.14; 13.76] | 0.32 [-0.34;0.98] |
| | Week 11 | 7.52 [5.75; 9.84] | 9.25 [7.08; 12.09] | 0.21 [-0.48;0.89] |
| | Week 12 | 6.93 [5.25; 9.16] | 9.93 [7.61; 12.97] | 0.39 [-0.35;1.10] |
| | (Week 13) | 8.96 [5.85;13.71] | 10.85 [7.74; 15.22] | 0.16 [-1.57;1.83] |
| | 1-month follow-up | 6.90 [5.29; 9.00] | 8.44 [6.44; 11.06] | 0.22 [-0.43;0.85] |
| | 6-month follow-up | 5.99 [4.54; 7.88] | 8.05 [6.12; 10.59] | 0.32 [-0.34;0.98] |
| PTCI | Baseline | 150.60 [135.73; 167.09] | 154.00 [138.03; 171.81] | |
| | 1-month follow-up | 106.05 [89.87; 125.14] | 130.28 [109.62; 154.82] | 0.21 [-0.76; 0.65] |
| | 6-month follow-up | 108.62 [90.52; 130.35] | 124.95 [103.23; 151.23] | 0.14 [-0.52; 0.79] |
| BDI-II | Baseline | 32.15 [27.94; 37.00] | 32.28 [27.83; 37.43] | |
| | 1-month follow-up | 21.15 [16.52; 27.08] | 25.83 [19.99; 33.39] | 0.22 [-0.42; 0.85] |
| | 6-month follow-up | 22.08 [16.89; 28.87] | 23.24 [17.53; 30.82] | 0.06 [-0.60; 0.71] |
| BAI | Baseline | 22.30 [17.52; 28.39] | 27.80 [21.52; 35.91] | |
| | 1-month follow-up | 16.40 [12.82; 20.98] | 20.22 [15.66; 26.12] | 0.24 [-0.40; 0.87] |
| | 6-month follow-up | 18.04 [14.07;23.13] | 20.97 [16.17; 27.20] | 0.18 [-0.48; 0.83] |

| | | | | |
|---|---|---|---|---|
| WHODAS | Baseline | 38.85 [34.38; 43.91] | 44.60 [39.21; 50.73] | |
| | 1-month follow-up | 28.75 [23.60; 35.02] | 41.32 [33.56; 50.87] | 0.73 [0.06; 1.37] |
| | 6-month follow-up | 28.67 [22.60; 36.37] | 36.79 [28.63; 47.29] | 0.42 [-0.25; 1.07] |
| GSE | Baseline | 22.80 [20.23; 25.69] | 20.50 [17.99; 23.36] | |
| | 1-month follow-up | 26.95 [24.06; 30.18] | 22.83 [20.13; 25.90] | -0.16 [-0.79; 0.48] |
| | 6-month follow-up | 27.19 [24.24; 30.49] | 22.62 [19.89; 25.71] | -0.18 [-0.83; 0.48] |
| PSQI | Baseline | 11.34 [9.79; 13.14] | 10.56 [9.03; 12.34] | |
| | 1-month follow-up | 7.77 [6.52; 9.25] | 8.03 [6.70; 9.63] | 0.03 [-0.62; 0.69] |
| | 6-month follow-up | 8.53 [7.12; 10.22] | 8.02 [6.62; 9.72] | -0.06 [-0.77; 0.64] |
| Sleep latency (stage 2), min | Baseline | 22.01 [14.88; 32.58] | 23.70 [15.43; 36.41] | -0.14 [-0.98; 0.70] |
| | 1-month follow-up | 19.65 [12.64; 30.55] | 24.06 [15.02; 38.55] | -0.33 [-1.21; 0.55] |
| Sleep efficiency, % | Baseline | 88.28 [84.71; 92.01] | 86.09 [82.28; 90.07] | 0.38 [-0.48; 1.21] |
| | 1-month follow-up | 89.96 [86.00; 94.09] | 86.93 [83.08; 90.96] | 0.48 [-0.42; 1.35] |
| Total sleep duration, min | Baseline | 425.48 [402.21; 450.09] | 401.94 [377.92; 427.48] | 0.62 [-0.26; 1.46] |
| | 1-month follow-up | 406.25[367.28; 449.35] | 396.94[358.52; 439.48] | 0.15 [-0.74; 1.02] |
| WASO duration, % | Baseline | 9.18 [6.11; 13.79] | 10.82 [6.92; 16.91] | -0.24 [-1.07; 0.61] |
| | 1-month follow-up | 7.37 [3.64; 14.93] | 9.37 [4.60; 19.11] | -0.27 [-1.14; 0.62] |
| Non-rem sleep (N1), % | Baseline | 10.32 [7.36; 14.46] | 8.07 [5.57; 11.68] | 0.41 [-0.45; 1.24] |
| | 1-month follow-up | 8.16 [5.73; 11.63] | 7.16 [4.95; 10.37] | 0.28 [-0.61; 1.15] |
| Non-rem sleep (N2), % | Baseline | 56.20 [52.34; 60.34] | 53.62 [49.56; 57.96] | 0.36 [-0.50; 1.19] |
| | 1-month follow-up | 57.53 [53.24; 62.16] | 56.03 [51.83; 60.57] | 0.27 [-0.62; 1.14] |
| Slow wave sleep (N3), % | Baseline | 14.97 [12.59; 17.80] | 20.24 [16.75; 24.47] | -0.97 [-1.82; -0.05] |
| | 1-month follow-up | 16.06 [13.39; 19.28] | 17.03 [14.09; 20.58] | -0.18 [-1.01; 0.67] |
| REM sleep, % | Baseline | 18.52 [14.87; 23.06] | 18.08 [14.21; 22.99] | 0.06 [-0.78; 0.90] |
| | 1-month follow-up | 21.48 [16.89; 27.31] | 17.86 [14.04; 22.72] | 0.55 [-0.37; 1.41] |
| REM density | Baseline | 16.58 [12.63; 21.78] | 20.20 [15.18; 26.88] | -0.17 [-1.00; 0.68] |
| | 1-month follow-up | 16.22 [13.28; 19.81] | 15.40 [12.50; 18.97] | 0.05 [-0.83; 0.92] |
| Movement arousal total sleep | Baseline | 88.83 [66.63; 118.44] | 77.90 [56.78; 106.88] | 0.12 [-0.73; 0.95] |
| | 1-month follow-up | 75.91 [56.01; 102.89] | 71.10 [51.78; 97.64] | 0.05 [-0.83; 0.92] |

*Abbreviations:* CAPS-5, Clinician-Administered PTSD Scale for DSM-5; PCL-5, Posttraumatic Stress Disorder Checklist, PTCI, Posttraumatic Cognitions Inventory; BDI-II, Becks Depression Inventory; BAI, Becks Anxiety Inventory; 'WHODAS 2.0, World Health Organization Disability Assessment Schedule 2.0; GSE, General Self-Efficacy Scale; PSQI, Pittsburgh Sleep Quality Index, WASO, wake after sleep onset, REM, rapid eye movement,
[1] SMD was calculated as the difference between transformed estimated mean scores of the treatment groups at each time point, divided by the pooled standard deviation (transformed scale) at each time point, based on a GLMM with gamma or negative binominal regression with only a fixed intercept.

**Time to Response, Remission, Recovery of self-rated PTSD symptoms regarding the index trauma**

The log-rank test revealed a statistically significant difference between remission rates over the duration of treatment sessions ($\chi^2$ (1) = 4.77, $p$ = .029). In the congruent condition, 17 participants experienced remission of PTSD symptoms regarding their index trauma (85%) compared with nine participants in the control condition (50%). Figure S3 shows the Kaplan-Meier survival curves for both conditions for the weekly treatment sessions. The estimated time to onset of remission within the treatment sessions was 4.45 weeks in the congruent condition (95% CI [2.50; 6.40]) and 8.11 weeks (95% CI [5.62; 10.60]) in the control condition. The unadjusted Cox regression showed a hazard ratio [HR] of 2.33 (95%CI [1.03; 5.27], $p$ = .043).

Regarding the response rates, the log-rank test revealed a statistically significant difference between response rates over the duration of treatment sessions ($\chi^2$ (1) = 6.36, $p$ = .012). In the congruent condition, 20 participants responded to treatment regarding their index trauma (100%) compared with 14 participants responding in the control condition (77,78%). Figure S2b shows the Kaplan-Meier survival curves for both conditions for the weekly treatment sessions. The estimated time to onset of response within the treatment sessions was 1.75 weeks in the congruent condition (95% CI [0.84: 2.66]) and 5.28 weeks (95% CI [2.83; 7.72]) in the control condition. The unadjusted Cox regression showed a hazard ratio [HR] of 2.28 (95%CI [1.07; 4.89], $p$ = .034).

Regarding the recovery rates, the log-rank test revealed a statistically significant difference between recovery rates over the duration of treatment sessions ($\chi^2$ (1) = 4.83, $p$ = .028). In the congruent condition, 13 participants recovered regarding their index trauma (65%) compared with 5 participants in the control condition (27.78%). Fig. S2c shows the Kaplan-Meier survival curves for both conditions for the weekly treatment sessions. The estimated time to onset of recovery within the treatment sessions was 7.80 weeks in the congruent condition (95% CI [5.81; 9.79]) and 10.78 weeks (95% CI [8.99; 12.56]) in the control condition. The unadjusted Cox regression showed a hazard ratio [HR] of 2.92 (95%CI [1.04; 8.21], $p$ = .043).

**Figure S3**. Kaplan–Meier estimates of time to treatment response and recovery by odour condition.

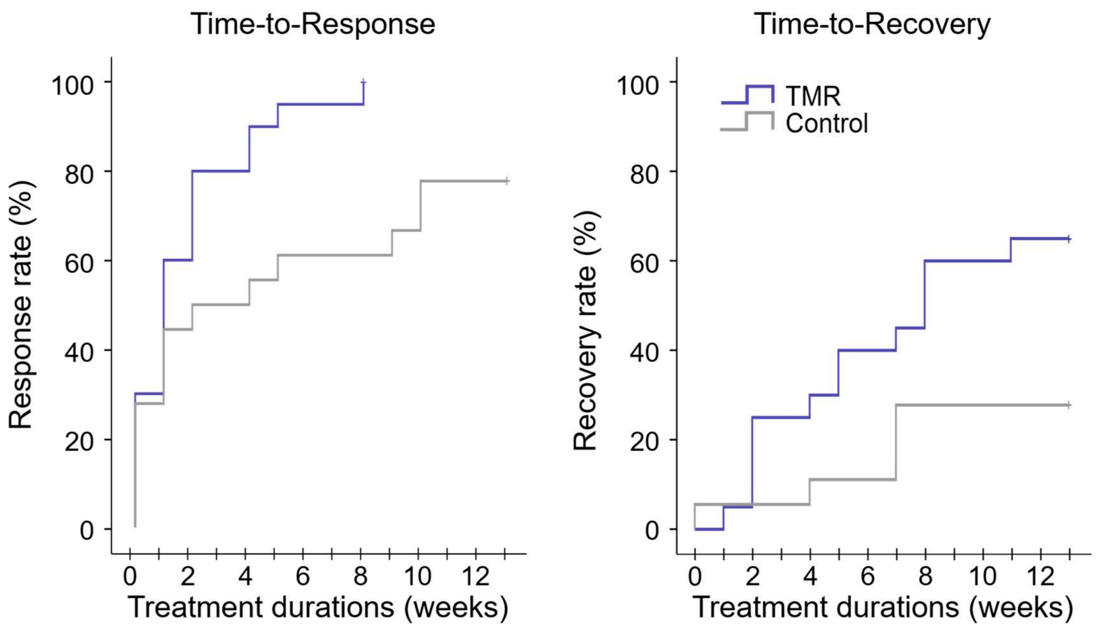

Curves show the cumulative proportion of patients meeting criteria for treatment response or recovery over time in the congruent (blue line) and the incongruent (grey line) condition. Response was defined as a reduction of ≥10 points from baseline on the PTSD Checklist for DSM-5 (PCL-5). Recovery was defined as a PCL-5 score ≤19 for the index trauma.

*Abbreviations:* TMR, Targeted memory reactivation: congruent-odour condition

**Table S8.** Minimal Clinically Important Difference (MCID) according to self-rated PTSD symptoms study 2

| | Total (*N* = 38) | Congruent (*n* = 20) | Incongruent (*n* = 18) | Congruent vs. Incongruent |
|---|---|---|---|---|
| Mid-treatment, *n (%)* | | | | $\chi^2 = 9.75$** |
| Improved | 23 (63.9) | 16 (88.9) | 7 (38.9) | |
| Not improved | 13 (36.1) | 2 (11.1) | 11 (61.1) | |
| Missing | 2 (5.3) | 2 (10.0) | 0 (0.0) | |
| One month Follow-up | | | | |
| Improved | 29 (76.3) | 16 (80.0) | 13 (72.2) | $\chi^2 = 0.32$ |
| Not improved | 9 (23.7) | 4 (20.0) | 5 (27.8) | |
| Deteriorated | 2 (5.3) | 1 (5.0) | 1 (5.6) | $\chi^2 = 0.01$ |
| Missing | 0 (0.0) | 0 (0.0) | 0 (0.0) | |

*Abbreviations:* PCL-5, Posttraumatic Stress Disorder Checklist 5, MCID was calculated using z-scores and a SD of 0.471, according to the recommendations of Stefanovics et. al.[1], * p < .05, ** p < .01, *** p < .001

**Table S9:** Test statistics of the repeated measures ANOVAs of the oscillatory EEG data in study 2

| | | Repeated Measures ANOVA* |
|---|---|---|
| Slow oscillations | Electrode*time*condition | $F(2, 15) = 0.69, p = .518$ |
| | Electrode*time | $F(2, 15) = 2.87, p = .088$ |
| | Time*condition | $F(1\ 16) = 0.43, p = .522$ |
| | Electrode*condition | $F(2, 15) = 0.21, p = .815$ |
| | Time | $F(1, 16) = 0.96, p = .342$ |
| Slow wave activity | Electrode*time*condition | $F(2, 15) = 0.89, p = .433$ |
| | Electrode*time | $F(2, 15) = 2.48, p = .118$ |
| | Time*condition | $F(1, 16) = 0.32, p = .580$ |
| | Electrode*condition | $F(2, 15) = 0.10, p = .902$ |
| | Time | $F(1, 16) = 0.23, p = .638$ |
| Fast spindle band | Electrode*time*condition | $F(2, 16) = 1.48, p = .156$ |
| | Electrode*time | $F(2, 16) = 0.61, p = .558$ |
| | Time*condition | $F(1, 17) = 0.10, p = .751$ |
| | Electrode*condition | $F(2, 16) = 1.44, p = .266$ |
| | Time | $F(1, 17) = 1.81, p = .196$ |

| | | | M (95% CI) | |
|---|---|---|---|---|
| | Electrode | Assessment | Congruent | Incongruent |
| Slow oscillations | Frontal electrode | Baseline | 35.69 [24.40; 46.98] | 40.20 [27.58; 52.82] |
| | | 1-month follow-up | 34.32 [22.81; 45.83] | 32.89 [20.03; 45.76] |
| | Central electrode | Baseline | 25.47 [15.50; 15.50] | 30.15 [19.00; 41.30] |
| | | 1-month follow-up | 25.34 [14.92; 35.75] | 26.79 [15.15; 38.44] |
| | Occipital electrode | Baseline | 10.18 [6.98; 13.39] | 12.15 [8.57; 15.73] |
| | | 1-month follow-up | 11.13 [4.93; 17.34] | 15.08 [8.14; 22.02] |
| Slow wave activity | Frontal electrode | Baseline | 13.40 [9.66; 17.15] | 14.67 [10.48; 18.86] |
| | | 1-month follow-up | 13.07 [9.17; 16.98] | 12.35 [7.99; 16.72] |
| | Central electrode | Baseline | 10.07 [6.74; 13.41] | 11.02 [7.29; 14.75] |
| | | 1-month follow-up | 10.01 [6.62; 13.40] | 10.13 [6.34; 13.92] |
| | Occipital electrode | Baseline | 3.83 [2.67; 4.99] | 4.22 [2.93; 5.51] |
| | | 1-month follow-up | 4.26 [2.186; 6.336] | 5.28 [2.96; 7.60] |
| Fast spindle band | Frontal electrode | Baseline | 0.27 [0.20; 0.34] | 0.21 [0.12; 0.29] |
| | | 1-month follow-up | 0.30 [0.23; 0.37] | 0.19 [0.11; 0.27] |
| | Central electrode | Baseline | 0.31 [0.23; 0.39] | 0.19 [0.10; 0.28] |
| | | 1-month follow-up | 0.32 [0.25; 0.39] | 0.20 [0.12; 0.28] |
| | Occipital electrode | Baseline | 0.13 [0.09; 0.17] | 0.09 [0.043; 0.14] |
| | | 1-month follow-up | 0.15 [0.09; 0.21] | 0.13 [0.05; 0.20] |

*Abbreviations:* EEG, electroencephalography, WASO, wake after sleep onset, REM, rapid eye movement, *2 time (baseline, FU) x 3 electrode (frontal, central, occipital) x 2 condition (congruent odour, incongruent odour), *log-transformed because of non-normally distributed data in a small sample, outliers (z-score >3 or <-3) were removed from the analyses; slow wave activity/ slow oscillations: $n_{congruent} = 10$, $n_{incongruent} = 8$; fast spindle band: $n_{congruent} = 11$, $n_{incongruent} = 8$

### Supplementary Methods

#### Deviations from the study protocol

*Sample size:* We adjusted the sample size calculation based on effect sizes of recent research in the field.

*Exclusion criteria study:* We expanded the exclusion criteria of study 2 according to common exclusion criteria in clinical trials and added a lifetime diagnosis of a psychotic or bipolar disorder, intellectual impairment, current substance use disorder and acute suicidality to the list of exclusion criteria

*Statistical analyses study 2:* We revised the statistical analysis plan to incorporate more up-to-date and sophisticated analytical methods that better fit the data structure and appropriately accounted for missing data.

Spectral analyses

All spectral analyses were conducted using custom MATLAB scripts based on the FieldTrip toolbox[2]. Analyses were restricted to artefact-free and arousal-free sleep epochs and performed separately for NREM (N2 and N3) and REM sleep.

To assess cueing-related oscillatory activity, EEG recordings were segmented into alternating 4-min odour ON and 4-min odour OFF periods. Following previous TMR studies, the first minute of each odour ON interval and the final minute of the preceding odour OFF interval were extracted for analysis. Data were divided into consecutive 8.192-s epochs with 50% overlap and multiplied by a Hanning window before spectral decomposition using fast Fourier transformation (FFT). Absolute power spectra were calculated between 0.125 and 45 Hz and averaged across epochs. Spectral power was subsequently averaged within predefined frequency bands: slow-wave activity (SWA; 0.5–4 Hz), slow oscillations (SO; 0.5–1.25 Hz), fast spindle activity (12–16 Hz), and REM theta activity (4.25–8 Hz).

Detection of slow oscillations

Slow oscillation detection followed established procedures[3]. EEG data were band-pass filtered between 0.3 and 3.5 Hz. Positive-to-negative zero crossings were identified and intervals shorter than 0.8 s or longer than 2 s were discarded, corresponding to oscillations in the 0.5–1.25 Hz frequency range.

For the remaining intervals, negative peak amplitudes and peak-to-peak amplitudes were calculated. Individual detection thresholds were derived separately for each electrode by multiplying the mean negative peak amplitude and mean peak-to-peak amplitude by 1.25. Events were classified as slow oscillations when both criteria were fulfilled: (i) the negative peak exceeded the amplitude threshold and (ii) the peak-to-peak amplitude exceeded the corresponding amplitude criterion.

Detection of sleep spindles

Sleep spindle detection was performed following previously validated procedures[3]. EEG data were band-pass filtered between 12 and 16 Hz. The root mean square (RMS) of the filtered signal was computed using a 200-ms sliding window and smoothed using an identical window length.

For each electrode, a detection threshold was defined as the mean RMS value across all NREM sleep epochs plus 1.28 standard deviations. Intervals during which the RMS signal exceeded this threshold for at least 0.4 s but not longer than 3 s were classified as spindle events.

SO–spindle coupling

SO–spindle coupling was quantified by determining whether the midpoint of a detected spindle event occurred within the time window of a detected slow oscillation, defined as the interval between two consecutive positive-to-negative zero crossings. The absolute number of coupled SO–spindle events was calculated separately for odour ON and OFF periods and used for subsequent statistical analyses.

Analyses of heart rate

ECG data recorded during script-driven imagery were processed using Kubios Heart Rate Variability (HRV) software (version 4.0[4]). R-waves were automatically detected using the built-in QRS detector algorithm based on the Pan–Tompkins method[6]. For preprocessing, ECG signals were band-pass filtered, squared, and smoothed using a moving average filter. R–R intervals were subsequently interpolated at 2000 Hz to improve temporal resolution. Noise segments were identified automatically based on both raw ECG signals and inter-beat interval (IBI) data and excluded from further analyses. Ectopic beats and artefacts were corrected using the automatic artefact correction procedures implemented in Kubios. The correction threshold was set to the medium level, identifying IBIs deviating by more than 0.25 s from the local average. Detected artefacts were replaced using cubic spline interpolation according to the procedures described by Tarvainen and Niskanen (2012)[4]. Heart rate responses were calculated separately for each phase of the script-driven imagery procedure. For statistical analyses, heart rate difference scores between negative and neutral memories were computed.

Randomization and masking

Study 1: The order of the congruent and incongruent cueing conditions was randomized across participants without stratification, given the relatively homogeneous sample and within-subject design. Participants were blinded to cueing condition throughout the study. Researchers involved in sleep scoring, EEG preprocessing, and electrophysiological analyses were also blinded to condition until completion of data analysis.

Study 2: Randomization was stratified by baseline PTSD severity, as assessed with the CAPS-5, to ensure balanced symptom severity across conditions. Participants were not blinded to the odour they received, as this was not feasible; however, they remained unaware of the nature of the experimental conditions and the study hypotheses. Baseline demographic and clinical characteristics were comparable between groups (Supplementary Table S5), indicating successful randomization. Research assistants, study coordinators, therapists, and clinical raters remained blinded to group assignment throughout the study.

Sample size calculation

Study 1: Sample size calculation was based on previous studies of olfactory TMR[8], which reported large effect sizes (Cohen's $d = 0.98$). Using an anticipated effect size of Cohen's $d = 1.0$, a statistical power of 0.8 and an alpha level of 0.05 resulted in a target sample size of N = 34.

Study 2: Sample size calculation was based on the expected between-group difference in PTSD symptom reduction. At the time of study planning, no previous studies had examined the effects of odour cueing as an adjunct to imagery rescripting for PTSD. Therefore, the calculation was informed by the largest available clinical TMR studies, including an odour-based TMR intervention conducted in a real-world setting (Cohen's $d = 1.20$)[9] and an auditory TMR intervention combined with imagery rehearsal therapy in patients with nightmare disorder ($d = 1.03$)[10]. Assuming an effect size of $d = 1.10$, a two-sided significance level of $\alpha = 0.05$, and a statistical power of 90%, a total sample of 38 completers (19 per group) was required. Allowing for an anticipated attrition rate of approximately 21%[11], the target sample size was set at 46 participants (23 per group).

Secondary outcomes

Study 2: Self-reported PTSD symptoms were assessed using the PTSD Checklist for DSM-5 (PCL-5)[12], a 20-item questionnaire assessing symptom severity during the previous week. Total scores range from 0 to 80, with higher scores indicating greater PTSD symptom severity. In addition to total scores, analyses were conducted for the corresponding DSM-5 symptom clusters. The PCL-5 has good psychometric properties and is sensitive to clinical change[12]. A score $\geq 31$ was considered indicative of probable PTSD, whereas a reduction of $\geq 10$ points from baseline was defined as clinically meaningful treatment response[13]. Remission was defined as a PCL-5 score $< 31$ and recovery as a score $\leq 19$, reflecting low end-state symptom levels and a low likelihood of ongoing PTSD diagnosis[14]. These criteria were also used for the time-to-event analyses of treatment response, remission, and recovery. Additional self-report outcomes included the Posttraumatic Cognitions Inventory (PTCI)[15], Beck Depression Inventory-II (BDI-II)[16], Beck Anxiety Inventory (BAI)[17], 32-item World Health Organization Disability Assessment Schedule 2.0 (WHODAS 2.0)[18], and General Self-Efficacy Scale

(GSE)[19]. The PCL-5 and PTCI were additionally assessed weekly throughout treatment to characterize symptom trajectories and trauma-related cognitions over time.

**References**

1. Stefanovics, E. A., Rosenheck, R. A., Jones, K. M., Huang, G. & Krystal, J. H. Minimal clinically important differences (MCID) in assessing outcomes of Post-Traumatic Stress Disorder. *Psychiatr. Q.* **89**, 141–155 (2018).
2. Oostenveld, R., Fries, P., Maris, E. & Schoffelen, J.-M. FieldTrip: Open source software for advanced analysis of MEG, EEG, and invasive electrophysiological data. *Comput. Intell. Neurosci.* **2011**, 156869 (2011).
3. Klinzing, J. G. *et al.* Auditory stimulation during sleep suppresses spike activity in benign epilepsy with centrotemporal spikes. *Cell Rep. Med.* **2**, 100432 (2021).
4. Tarvainen, M. P., Niskanen, J.-P., Lipponen, J. A., Ranta-Aho, P. O. & Karjalainen, P. A. Kubios HRV--heart rate variability analysis software. *Comput. Methods Programs Biomed.* **113**, 210–220 (2014).
5. Pan, J. & Tompkins, W. J. A real-time QRS detection algorithm. *IEEE Trans. Biomed. Eng.* **32**, 230–236 (1985).
6. Lieber, T. Targeted Memory Reactivation increases memory recall: A meta-analysis. *bioRxiv* (2019) doi:10.1101/796458.
7. Neumann, F., Oberhauser, V. & Kornmeier, J. How odor cues help to optimize learning during sleep in a real life-setting. *Sci. Rep.* **10**, 1227 (2020).
8. Schwartz, S., Clerget, A. & Perogamvros, L. Enhancing imagery rehearsal therapy for nightmares with targeted memory reactivation. *Curr. Biol.* **32**, 4808-4816.e4 (2022).
9. Varker, T. *et al.* Dropout from guideline-recommended psychological treatments for posttraumatic stress disorder: A systematic review and meta-analysis. *J. Affect. Disord. Rep.* **4**, 100093 (2021).
10. Weathers, F. W., Litz, B. T., Keane, T. M., Palmieri, P. A. & Marx, B. P. The ptsd checklist for dsm-5 (pcl-5). (2013).
11. Forkus, S. R. *et al.* The posttraumatic stress disorder (PTSD) Checklist for DSM-5: A systematic review of existing psychometric evidence. *Clin. Psychol. (New York)* **30**, 110–121 (2023).
12. Blevins, C. A., Weathers, F. W., Davis, M. T., Witte, T. K. & Domino, J. L. The Posttraumatic Stress Disorder Checklist for DSM-5 (PCL-5): Development and initial psychometric evaluation: Posttraumatic stress disorder checklist for DSM-5. *J. Trauma. Stress* **28**, 489–498 (2015).
13. Foa, E. B., Ehlers, A., Clark, D. M., Tolin, D. F. & Orsillo, S. M. The Posttraumatic Cognitions Inventory (PTCI): Development and validation. *Psychol. Assess.* **11**, 303–314 (1999).
14. Inventory-Ii, B. D. Beck depression inventory-II. *Corsini Encycl. Psychol* **1**, 210 (2010).
15. Beck, A. T. & Steer, R. A. Beck anxiety inventory (BAI).
16. Üstün, T. B. *et al.* Developing the World Health Organization disability assessment schedule 2.0. *Bull. World Health Organ.* **88**, 815–823 (2010).
17. Luszczynska, A., Scholz, U. & Schwarzer, R. The general self-efficacy scale: multicultural validation studies. *J. Psychol.* **139**, 439–457 (2005).
18. Smyth, C. The Pittsburgh sleep quality index (PSQI). *J. Gerontol. Nurs.* **25**, 10–11 (1999).
19. Moher, D. *et al.* CONSORT 2010 explanation and elaboration: updated guidelines for reporting parallel group randomised trials. *Int. J. Surg.* **10**, 28–55 (2012).
